# Flexible and Stable Cycle-by-Cycle Phase-Locked Deep Brain Stimulation System Targeting Brain Oscillations in the Management of Movement Disorders

**DOI:** 10.1101/2025.04.16.25325658

**Authors:** Xuanjun Guo, Alek Pogosyan, Jean Debarros, Shenghong He, Laura Wehmeyer, Fernando Rodriguez Plazas, Karen Wendt, Zixiao Yin, Ahmed Raslan, Thomas Hart, Francesca Morgante, Tim Denison, Erlick A Pereira, Keyoumars Ashkan, Shouyan Wang, Huiling Tan

**Affiliations:** Institute of Science and Technology for Brain-Inspired Intelligence, Fudan University, Shanghai, China; Medical Research Council Brain Network Dynamics Unit, Nuffield Department of Clinical Neurosciences, University of Oxford, Oxford, United Kingdom; MOE Frontiers Center for Brain Science, Fudan University, Shanghai, China; Department of Engineering Science, University of Oxford, Oxford, United Kingdom; Department of Neurosurgery, Beijing Tiantan Hospital, Capital Medical University, Beijing, China; Department of Neurosurgery, King’s College Hospital, Denmark Hill, London, United Kingdom; City St George’s, University of London & St George’s University Hospitals NHS Foundation Trust, Neuroscience & Cell Biology Research Institute, Cranmer Terrace, London, United Kingdom

**Keywords:** Phase-locked stimulation, Kalman filter, Non-resonant oscillators, Parkinson’s disease, Closed-loop neuromodulation

## Abstract

**Background:** Phase-locked neuromodulation aligns electrical or magnetic stimulation with the brain’s natural rhythms, showing promising potential to enhance therapeutic outcomes by more precisely modulating specific neural oscillations. However, stimulation-induced artifacts critically compromise real-time phase estimation accuracy. Existing approaches either suspend phase-tracking following stimulation or employ dedicated hardware systems yet introduce estimation instability through temporal gaps and signal distortion.

**Objective:** We develop and evaluate a flexible and stable phase-locked deep brain stimulation (PLDBS) system capable of delivering cycle-by-cycle phase-aligned stimulation based on brain oscillations, with an additional focus on its potential for modulating movement.

**Methods:** The PLDBS system was implemented using portable CE-marked devices and a computer-in-the-loop framework. Simulations and clinical experiments were performed targeting distinct phases of neural oscillations. The simulation framework evaluated the real-time performance of different phase-tracking methodologies considering artifacts, ultimately establishing a Kalman filter-based artifact removal system integrated with non-resonant oscillators for instantaneous phase estimation, thereby defining the final cycle-by-cycle PLDBS architecture. We then evaluated the performance of the pipeline for PLDBS in human patients targeting cortical alpha and subthalamic nucleus (STN) beta rhythms.

**Results:** Our system achieved over 90% accuracy in delivering stimulation within a 90°and 45°window centered around the target phase for STN beta (proximal recording) and cortical alpha rhythms (distal recording), respectively. Stimulation delivered at different STN beta phases led to a significant difference in evoked potentials in STN local field potentials in 3 out of 4 participants. However, such an effect was not found in cortical alpha in any participants. STN beta-triggered stimulation showed potential phase-dependent modulation of finger-tapping velocity and amplitude in Parkinson’s disease.

**Conclusion:** This study presents a flexible and stable pipeline for precise PLDBS with CE-marked devices and a computer-in-the-loop. Using this pipeline, we showed that PLDBS at different STN beta phases differentially modulates the evoked action potentials in the STN and motor behavior used to quantify bradykinesia, paving the way for further studies and clinical trials for PLDBS.

## Introduction

Phase-locked neurostimulation ^1,2^ is an innovative neuromodulation technique that harnesses the principles of synchronizing stimulation and neuronal oscillations to modulate brain activity for therapeutic purposes ^3–7^. Phases of a neural oscillation indicate moment-to-moment fluctuations in neuronal excitability ^8–10^, suggesting that stimulation delivered at specific phases can lead to more precise modulation of neuron population activities and, consequently, more pronounced physiological and behavioral effects ^11–20^. Targeting particular phases of neural oscillations with different stimulation modes, including transcranial magnetic stimulation (TMS)^21^, transcranial electrical stimulation^16,22,23^, deep brain stimulation (DBS)^2,24,25^, and acoustic stimulation^26,27^, has shown potential in influencing neuronal rhythms and improving cognition^5,28,29^, movement^30^ and sleep^22,26,27^. These approaches show promise for therapeutic applications in disorders such as essential tremor^13,31^, Parkinson’s disease (PD)^32,33^, and depression^34,35^, by precisely modulating brain network dynamics, which can subsequently influence behavior. While preclinical studies in movement disorders have indicated the efficacy of phase-targeted neuromodulation in bidirectionally regulating oscillatory dynamics (particularly subthalamic nucleus (STN) beta oscillations ^32,33^) to potentially modify motor outcomes^36^, critical gaps persist in translating these electrophysiological interventions into clinically viable behavioral modulation paradigms.

Achieving effective phase-locked neurostimulation requires overcoming two interdependent technical barriers: robust elimination of stimulation artifacts that obscure endogenous neural oscillations during stimulation and precise tracking of instantaneous phase dynamics^37^. Various methods have been proposed for artifact mitigation including bandpass filter^38–40^, template subtraction^24,25,41^, and blanking combined with interpolation^32,42–44^. In parallel, techniques such as phase inversion detection^45,46^, phase-locked oscillators^14,47^, analytic signal construction^23,48,49^, state-space model^32,50,51^ and resonant theory^52^ have been employed for real-time phase estimation. Despite these advances, cycle-by-cycle phase-locked DBS (PLDBS) targeting neural signals near stimulation sites still faces challenges^53^. Specifically, stable device-independent artifact removal methods remain elusive, and optimal approaches for accurate phase estimation in the presence of residual artifacts have yet to be established.

This study proposes a flexible and stable PLDBS pipeline to address these challenges that integrates an effective artifact removal method with an optimized real-time phase estimation algorithm, ensuring reliable cycle-by-cycle phase tracking. This pipeline is compatible with commonly used amplifiers. Using a computer-in-the-loop for real-time signal processing, we evaluated its real-time phase estimation accuracy and stability in cycle-by-cycle PLDBS targeting cortical and subthalamic oscillations in human patients. Our findings establish a methodological foundation for the clinical translation of phase-specific neuromodulation. Moreover, our results suggest that PLDBS at different STN beta phases differentially modulates evoked resonant neural activity (ERNA) in the STN, a modulation-responsive electrophysiological biomarker reflecting basal ganglia-thalamocortical circuit reorganization in PD^54–56^, and motor behavior related to bradykinesia, underscoring its potential for clinical application and paving the way for further studies and clinical trials.

## Material and Methods

### Real-time artifact removal method

We first reviewed several stimulation artifact removal methods, including template subtraction, interpolation, and sampling, as described in Supplementary materials (Supp Table 1).

**Table 1.** Clinical information.

|  | Gender | Diagnose | Handedness | DBS system | Cortical Alpha | STN beta |
| --- | --- | --- | --- | --- | --- | --- |
| 1 | Male | CD | Left | Boston | 10Hz | 25Hz |
| 2 | Male | PD | Left | Medtronic | 6Hz | 15Hz |
| 3 | Female | PD(Tremor) | Right | Medtronic | 9Hz | 18Hz |
| 4 | Male | PD(Bradykinesia) | Right | Medtronic | 10Hz | 19Hz |

Note: Participant 2 exhibited dominant 6Hz theta activity during eyes-closed across all EEG channels, prompting theta-band ( $6 \pm 1.5$ Hz) PLDBS implementation; Participant 3 demonstrated prominent 7 Hz resting tremor with bi-frequency alpha peaks (7Hz/9Hz), requiring selective 9 Hz alpha targeting. Abbreviation: CD: Cervical dystonia, PD: Parkinson's disease, DBS: Deep brain stimulation, STN: Subthalamic nucleus

Template subtraction assumes that the artifacts associated with each stimulation pulse follow a constant pattern. However, this assumption only applies when amplifiers with very high-sampling-rate (e.g. 44k Hz) and large dynamic ranges. In most cases, stimulation artifacts appear as random waveforms^41^, making template subtraction ineffective. Beyond that, recently proposed methods such as blanking or irregular sampling^43^ and interpolation depend on precisely knowing the stimulation artifact duration. The high-frequency components of square-waved stimulation pulses cause prolonged signal ringing after amplifier anti-aliasing filtering, representing its “impulse” response to the stimulation pulse. Thus, the type of frontend circuit, the analog-to-digital converter (ADC) technology, and the digital filtering technique influence the stimulation artifact duration and temporal shape, making removing it extremely challenging to mitigate with standard methods ^57^.

The Kalman filter ^58,59^ reduces uncertainty in the estimated signal by combining direct measurements with estimations based on a model, following Bayesian principles. It employs a “blanking” mechanism, where the confidence in the measurements is set low when stimulation artifacts are present. During these periods, the signal estimation is instead driven by a model. This study proposes using the Kalman filter for artifact removal with a second-order autoregressive (AR) model based on past data points. Unlike simpler interpolation techniques that either skip or replace artifacts with preset values, the Kalman filter adapts to variations in the signal, providing a more precise reconstruction of underlying neural activity.

To assess the Kalman filter’s artifact suppression stability, we compared three conditions: raw signals, artifact-contaminated signals, and Kalman-processed signals (Supp Fig 4). The Kalman filter outperformed irregular sampling methods under matched parameters, particularly in scenarios involving stimulation duration underestimation (Supp Table 3). This is a critical PLDBS challenge, as system-specific jitter and delay randomness can impede precise artifact landmark detection.

### Real-time phase estimation method

Several methods for phase estimation proposed in previous studies were considered, including zero-crossing (ZC), phase-locked oscillators, causal Hilbert transform (HT) methods such as endpoint-corrected HT (ecHT) ^49^ and autoregressive HT (arHT) ^15,48,60,61^, state space phase estimator (SSPE) ^50^, resonant oscillators (RO), non-resonant oscillators (NRO) ^52,62^ and OscillTrack ^32^, as detailed in Supplementary materials (Supp Table 2). Some of these approaches have been compared in previous literature (Supp Fig 1) ^50–52,62^. NRO has been shown to outperform RO, ecHT, and phase-locked oscillators in neural oscillations, offering a broader targeted frequency band. SSPE also outperforms the HT and ZC approaches, showing greater tolerance to noise. However, no study has identified a stable method for PLDBS scenarios, especially when integrated with stimulation artifact removal methods.

We implemented zero-crossing (ZC), autoregressive HT (arHT), non-resonant oscillators (NRO), state space phase estimator (SSPE), and OscillTrack in our study based on prior research. We compared their performance in a simulation model using real recordings and synthetic signals containing stimulation artifacts.

### Simulink model

To identify a stable method for phase estimation under varying signal-to-noise ratios and stimulation artifacts, we implemented and tested the different methods using Simulink, a MATLAB-based modeling environment. The designed model incorporates several modules, including data input, artifact removal, preprocessing, phase estimation, and stimulation (adding stimulation artifacts). After initial artifact suppression, the signals undergo fourth-order Chebyshev Type I bandpass filtering to ensure narrow-band frequency isolation. Although the IIR bandpass filter introduces a non-linear phase response across frequency bands (Supp Fig 2), it maintains an approximately linear phase response within the passband. It preserves most amplitude information, enabling the phase estimation method to retrieve the real-time phase (Supp Fig 3) effectively. The conditioned signals are then processed through the mentioned phase tracking algorithms to compute instantaneous phase dynamics. Finally, the model triggers phase-locked stimulation, where stimulation pulses are delivered at the target phase, with the stimulation artifacts added to the original input signal (Fig 1B). The methodological configurations for phase estimation were derived from the previously proposed optimization framework ^15,48,60,61^.

**Fig 1.**
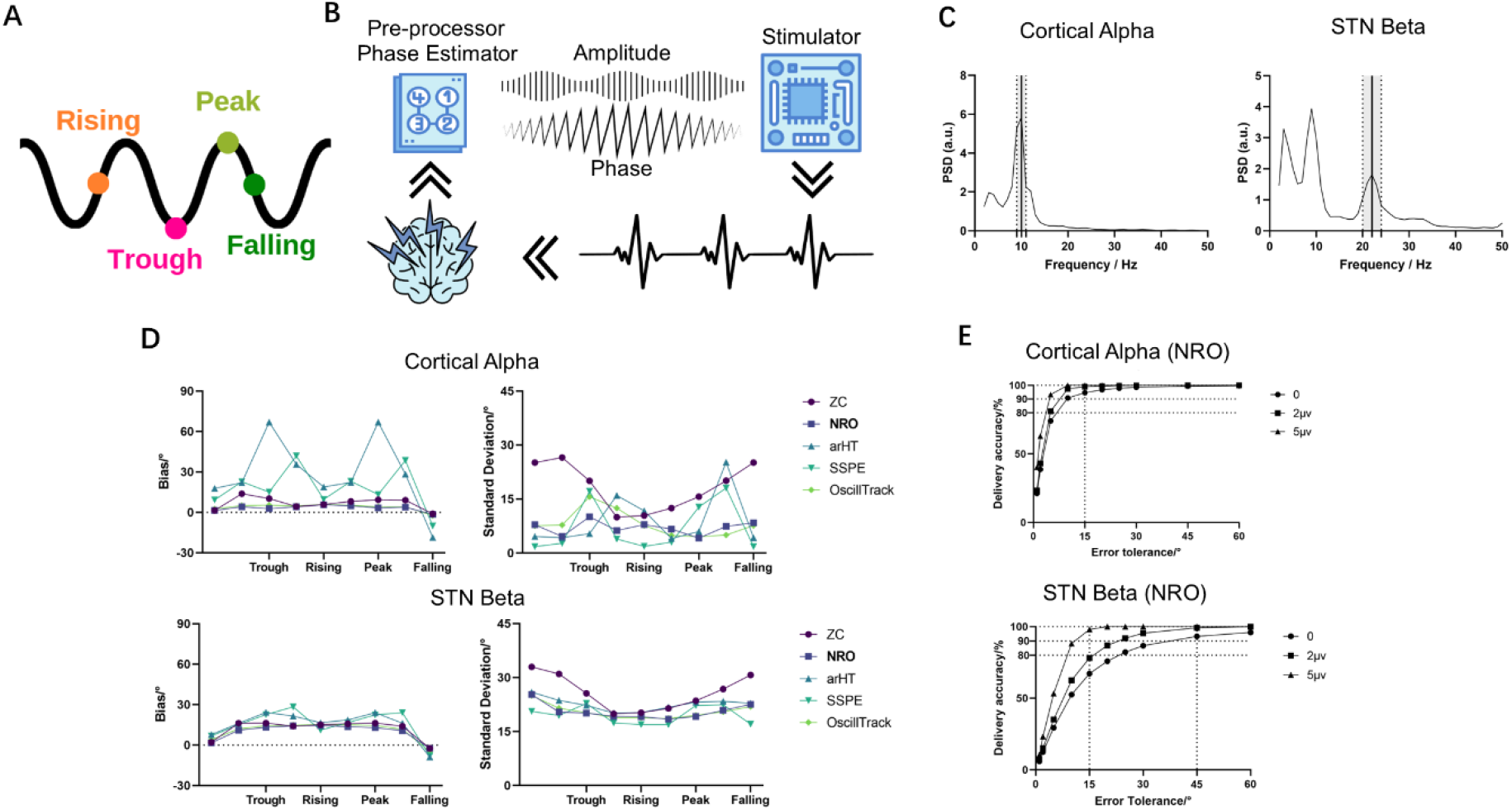
Performance evaluation of PLDBS using Kalman filtering and NRO analysis. (A) Schematic illustration of phase definitions: rising (0°), peak (90°), falling (180°), and trough (-90°). (B) Simulation of real-time PLDBS system. Neural signals undergo three-stage processing: (1) Artifact removal via Kalman filtering with bandpass filtering, (2) Instantaneous phase estimation through NRO, and (3) Phase-triggered stimulation delivery. (C) Spectral characteristics of cortical alpha and STN beta oscillations. Peak frequencies (vertical bars) with corresponding filter bandwidths (dashed lines) demonstrate oscillation-specific spectral filtering. (D) Phase estimation performance comparison across five methods. NRO showed superior performance with minimal *bias* and reduced *std*. (E) Stimulation timing accuracy showed precise targeting simulation with cortical alpha and STN beta oscillations. Abbreviation: PLDBS: phase-locked deep brain stimulation, PSD: power spectral density, STN: subthalamic nucleus, ZC: Zero-crossing, NRO: Non-resonant oscillators, arHT: autoregressive Hilbert Transform, SSPE: state space phase estimator

### Simulation input data and validation

Validation was performed using real recordings. The first dataset comprised 25-second eyes-closed resting-state electroencephalogram (EEG) recordings from the Pz electrode in a healthy participant, with robust cortical alpha oscillations (Fig 1C, left). The second dataset consisted of 60-second STN LFP recordings from a PD patient during OFF medication and OFF stimulation, exhibiting pathologically elevated beta-band synchrony (Fig 1B, right), derived from our previously published paper^63^. Both clinical datasets were sampled at 4096Hz and preprocessed through identical 2 Hz fourth-order Butterworth high-pass filtering. The local ethics committees approved the study, and all patients provided written informed consent according to the Declaration of Helsinki.

For each dataset, the instantaneous phase of the target oscillation was computed offline using the “gold standard” technique (i.e., offline Hilbert transform (oHT)), applied to the raw signal without artifacts. To systematically evaluate the phase-locking performance of different algorithms, we implemented a simulation framework that spanned the entire phase space (-180°to 180°, 45° increments). Two key metrics were used for evaluation: *bias* (absolute mean error between real-time delivered phases and offline gold-standard target phases) and the *std* (standard deviation of delivered phases) during each phase-locked stimulation simulation.

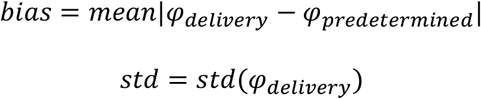

Further quantification included phase-specific *accuracy*, defined as the proportion of pulses delivered within incremental tolerance thresholds (0°, 2°, 5°, 10°, 15°, 20°, 25°, 30°, 45°, 60°), and calculated as:

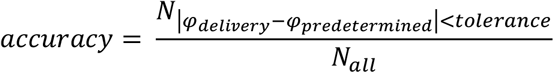

Since estimation errors may vary with the amplitude of the target oscillation^52^, amplitude-dependent gating thresholds of 2 µV and 5 µV were applied to the offline signal. The accuracy of the signal segments above these thresholds was evaluated to determine potential improvements in stimulation *accuracy* .

### PLDBS hardware implementation

Following computational validation, we implemented a closed-loop PLDBS system integrating CE-marked amplifiers (TMSi Saga, TMS International, Netherlands) and CE-marked stimulators (ISIS, Inomed Neurocare Ltd., Germany) with real-time processing via MATLAB and Lab Streaming Layer (Fig2A/4A). Signals (4096Hz unipolar sampling) underwent 2 Hz Butterworth high-pass filtering before Kalman filter-based artifact suppression and NRO phase estimation. Once the difference between the estimated and target phases drops below a specific threshold (set as 2°), a trigger signal is sent to the neurostimulator via Labjack. This brief TTL pulse trigger signal enables the stimulator to deliver a single stimulation pulse.

**Fig 2.**
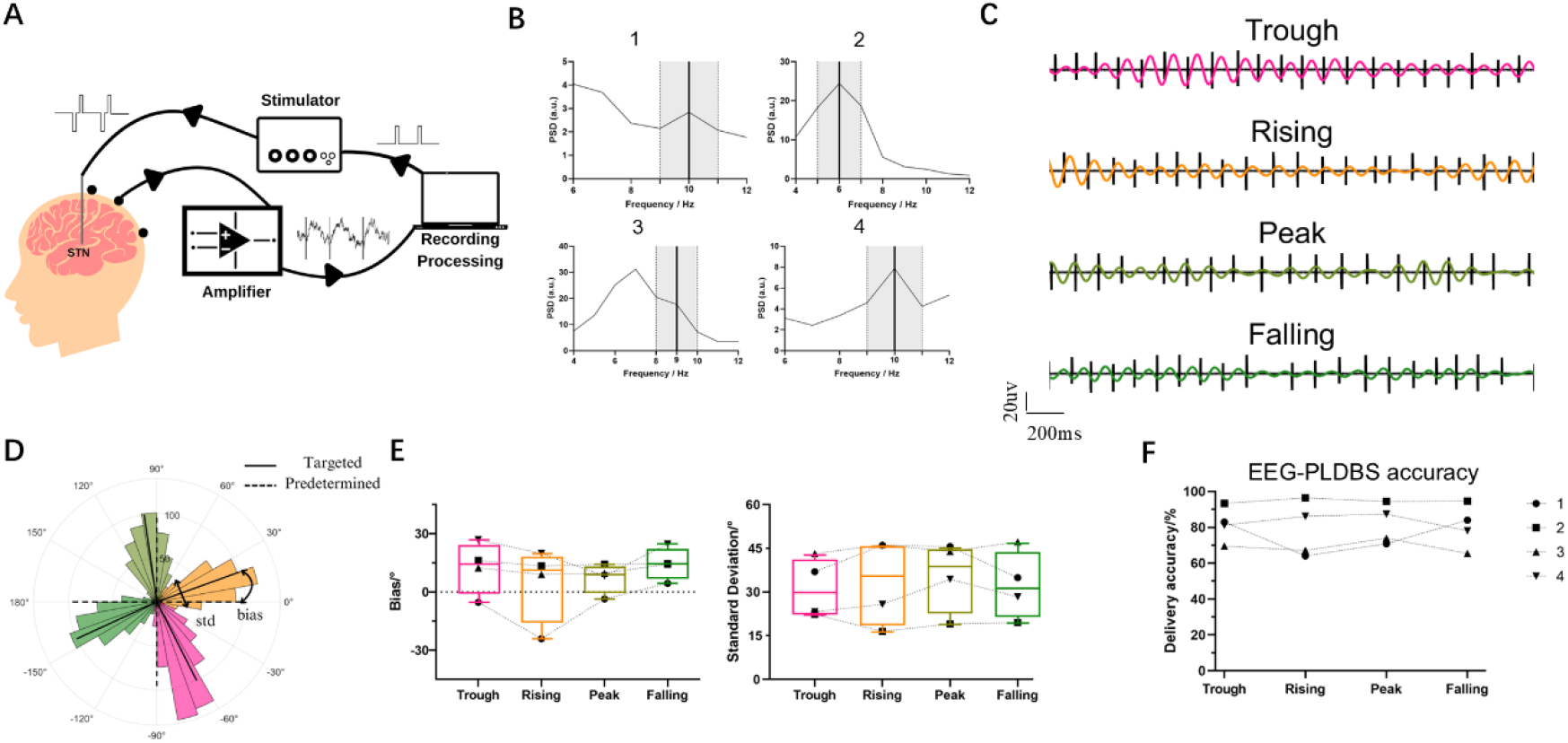
PLDBS of cortical alpha oscillations. (A) The PLDBS system delivered electrical pulses to the STN based on alpha oscillations recorded from an EEG channel. (B) Individualized spectral profiles of the four patients. Vertical lines denote individualized alpha peak frequency with 3 Hz bandwidth filters (gray shaded area). (C) Phase-locked stimulation exemplar (Participant 4, Oz channel, left hemisphere targeting). Representative 2-second traces show stimulation timing at four quadrant alpha phases: trough (pink), rising (orange), peak (yellow-green), and falling (dark green). Wideband EEG with stimulation artifacts is shown in black. (D) Angular distribution of stimulation phases for Participant 4. Circular histograms show phase concentration at targets (dashed lines) with mean vectors (black lines). (E) Phase estimation consistency across participants. Mixed-effects ANOVA revealed no significant phase-dependent *bias* (left) or *std* (right). (F) Targeting accuracy across phase quadrants. Overall system achieved 81.24% ± 3.53% accuracy (±45°tolerance) with comparable performance across phases.

Artifact detection is triggered at a 300 µV threshold. Based on previously recorded data ^63^ and analysis results in supplementary materials (Supp Table 3, Fig 4), the Kalman filter estimates LFPs during the subsequent 9 ms. For real-time phase estimation, the NRO method, which outperformed others in simulations, was implemented. This method uses a linear oscillator model 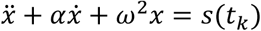, solved numerically to map the analytic input signal’s oscillator dynamics (amplitude/phase). Parameters (*α*_*a*_ = 80, *α*_*p*_ = 10, ω = 5ϑ, ϑ = 2π · *f*_*peak*_) followed methodology in ^64^, with *f*_*peak*_ as target oscillation frequency.

### PLDBS test in human patients

The PLDBS system was tested in four patients (Table 1), including one with cervical dystonia and three with PD, all undergoing STN targeted DBS surgery. The implanted DBS systems were from two manufacturers: Medtronic Inc. (USA) with octopolar directional leads (SenSight™ model 33005), and Boston Scientific (USA) with octopolar directional leads (Vercise™ model DB-2202). Electrodes were connected to temporary lead extensions and externalized through the temporal or frontal scalp.

Recordings were performed in the OFF-dopaminergic medication state, four to six days postoperatively. Monopolar stimulation was applied using an ISIS neurostimulator, with a self-adhesive electrode placed on the patient’s back as the reference. The stimuli consisted of symmetric, constant-current, biphasic pulses (3 mA, 60 µs, with the negative phase delivered first). The local ethics committees approved the study, and all patients provided written informed consent according to the Declaration of Helsinki.

Patients remained in a resting seated state throughout the entire recording session. Pre-experimental contact optimization involved 25-pulse mapping (4 mA, 1 ∼ 0.5 Hz) across STN channels, selecting contacts that elicited maximal evoked resonant neural activity (ERNA) amplitudes in adjacent recordings. A 10-second calibration block was used to quantify fixed system delay (median value of time intervals between trigger onset and stimulation artifact appearance in LFP traces) and temporal jitter (the range of delay measurements across all pulses), which remained constant through all experiments. Multimodal recordings (STN-LFP and EEG, bilateral fingertip accelerometry) were synchronized throughout phase-locked protocols.

### Cortical alpha-triggered PLDBS

Alpha oscillations are commonly observed in the parietal lobes via EEG recordings, especially with eyes closed, and are associated with rhythmic fluctuations in excitability^65,66^. To validate the PLDBS efficacy, we implemented cortical alpha-triggered PLDBS as a proof-of-concept demonstration (Fig 2A). Before stimulation, participants were asked to close their eyes while feedback channels exhibiting the highest alpha-band spectral power were selected from standard EEG (Fz, Cz, Pz, Oz). Individualized alpha peak frequencies were identified using power spectra density (PSD) analysis and extracted with 3 Hz bandwidth filters centered on the dominant frequency (Fig 2B). The cortical alpha-triggered PLDBS protocol targeted four cardinal phase points: rising (0°), peak (90°), falling (180°), and trough (-90°) see as Fig 1A. Each phase condition was tested in duplicate 30-second stimulation trials (8 total blocks) administered in a randomized sequence, with 60-second inter-block washout intervals.

### STN beta-triggered PLDBS

Beta oscillations in STN have been associated with bradykinesia and rigidity in PD, and STN beta-triggered PLDBS may more effectively modulate these pathological rhythms^67–69^. We implemented STN beta-triggered PLDBS in four participants (Fig 4A). The feedback channel was selected based on the contact exhibiting the highest ERNA amplitude during contact optimization. Participant-specific beta peak frequencies were derived from resting-state PSD analyses (OFF stimulation) and extracted using a 5Hz bandwidth filter (Fig 4B). Consistent with prior studies ^36,70^, STN beta-triggered PLDBS targeted two antithetical oscillation phases: rising (0°or 20°) and falling (180°or -160°). Each phase condition underwent triplicate 30-second stimulation trials (6 total blocks), administered in a randomized sequence, with 60-second inter-block washout intervals. Because the dual-phase paradigm is constrained by system latency parameters (∼10 ms jitter), corresponding to 30∼50% of beta-cycle durations (18-30Hz, 33-55ms/cycle), achieving higher phase resolution within the beta band was not feasible.

### In-vivo validation

Post hoc signal processing employed the same Kalman filter for artifact removal and used oHT as the golden standard to assess PLDBS targeting precision. Performance was evaluated using the same metrics as in simulations: *bias* and *std*. Phase-specific *accuracy* was quantified in two ways: per-target-phase participant means and population-level cross-participant averages. Different phase-locking tolerances were applied based on the oscillation type (cortical alpha-triggered-PLDBS: ±45°; STN beta-triggered PLDBS: ±90°). Consistent with the simulation approach, amplitude-gating thresholds were used to assess the effect of signal amplitude on *accuracy* .

### ERNA modulation with PLDBS

Stimulation phases may bidirectionally modulate beta oscillation amplitudes ^32,33,71^, but whether these effects relate to stimulation artifacts remains unresolved. Thus, we investigated phase-dependent modulation of ERNA (a candidate electrophysiological biomarker for neuromodulation treatment efficacy, mechanistically linked to STN neuronal firing dynamics ^54^) and implemented a standardized feature extraction pipeline based on established frameworks ^56,72^. ERNA waveforms were quantified using a custom MATLAB algorithm implementing multiscale peak detection. Signals containing less than two definable peak-trough pairs were excluded via automated quality control. ERNA amplitude was quantified as the absolute difference between the first positive peak and subsequent negative trough. Per-block ERNA amplitudes were calculated as the mean of valid pulse responses occurring within ±45°and ±90°tolerance in cortical alpha-triggered PLDBS and ±90°tolerance in STN beta-triggered PLDBS.

### Finger-tapping task with STN beta-triggered PLDBS

Participant 4, who showed predominant bradykinesia and rigidity, completed a finger-tapping protocol to assess the potential motor modulation effects of PLDBS. As an active control, 130Hz continuous DBS (cDBS) was applied first in two 15-second trials, with OFF recordings before and after serving as baselines (Fig6A Top). The PLDBS protocol consisted of two OFF-control blocks and six phase-locked stimulation blocks (Fig 6A Bottom), each separated by a 3-minute washout. Control blocks included pre- and post-15-second trials, while stimulation blocks delivered 60-second trains followed by 15-second finger-tapping under active PLDBS. Phase targeting alternated rising and falling oscillation phases (3 trials each) in a randomized order. The kinematic analysis focused on the axis of maximal acceleration amplitude, considering only accelerometer-discernible tapping peaks. Velocity was derived from the reciprocal inter-peak intervals (1/*Δt*), while tapping amplitude was quantified by peak-to-peak accelerometric magnitude.

### Statistics

PLDBS feasibility was assessed as follows: (1) Cortical alpha-phase analysis: Ordinary one-way ANOVA to compare *bias* and *std* across four phases; (2) STN beta-phase performance: Unpaired t-tests to compare rising vs falling phase; (3) Stimulation error-amplitude correlations: Pearson’s correlation analysis; (4) Amplitude-gating efficacy: Two-way ANOVA (phase×threshold) with multiple comparisons. (5) ERNA modulation: Ordinary one-way ANOVA for cortical alpha-triggered PLDBS across four phases and unpaired t-tests for both cortical alpha-triggered PLDBS and STN beta-triggered PLDBS (rising vs falling). (6) Movement modulation: Unpaired t-tests comparing cDBS ON/OFF and ordinary one-way ANOVA with multiple comparisons for PLDBS phases (rising/falling/OFF).

All analyses were conducted using custom MATLAB scripts. Correlations are reported as Pearson’s R (95% confidence interval), and all data are presented as *mean* ± *SD*. Post hoc multiple comparisons were conducted using Tukey’s HSD procedure with a family-wise error rate controlled at *α* = 0.05.

## Results

### NRO phase estimation following Kalman filter artifact removal demonstrates stability

Fig 1B illustrates the PLDBS simulation pipeline using real recordings, where recorded neural signals were processed in real-time for oscillation phase/amplitude estimation, followed by phase-triggered stimulation. Comparative analysis of estimation *bias* across different methods against oHT (Cortical alpha: ZC: 6.81°± 4.62°, NRO: 3.18°± 6.12°, arHT: 28.93°± 26.25°, SSPE:18.14°± 15.84°, OscillTrack: 3.88°± 2.52°; STN Beta: ZC: 11.99°± 7.00°, NRO: 9.55°± 6.12°, arHT: 15.16°± 10.32°, SSPE:15.45°± 11.11°, OscillTrack:10.78°± 6.84°) identified NRO and OscillTrack as the most accurate (Fig 1D, Left). For stability metrics (Fig 1D, Right), NRO and SSPE demonstrated superior artifact robustness, exhibiting lower *std* (Cortical alpha: ZC: 18.37°± 6.50°, NRO: 7.02°± 1.86°, arHT: 9.05°± 7.34°, SSPE:7.00°± 6.93°. OscillTrack:8.13°± 3.69°; STN beta: ZC: 25.80°± 4.91°, NRO: 20.58°± 2.14°, arHT: 22.60°± 1.79°, SSPE:19.53°± 2.55°. OscillTrack:20.53°± 2.04°). Given its optimal phase-tracking precision and artifact-resistant stability, NRO was selected as the final phase estimation method for PLDBS implementation.

Stimulation *accuracy* in PLDBS using the Kalman filter and NRO was evaluated next. For cortical alpha oscillations, stimulation *accuracy* reached 94.63% within a ±15°tolerance. Similarly, for STN beta oscillations, *accuracy* was 93.24% within a ±45°tolerance (Fig 1E). Notably, when the amplitude of the target oscillation exceeds 2 µV, STN-beta-triggered PLDBS achieved an even higher *accuracy* of 95.42% within a ±30°tolerance.

### Real-time EEG-triggered PLDBS for precise alpha phase targeting

Following a bench test using constant 20 Hz oscillations to validate the system, we conducted cortical alpha-triggered PLDBS in a clinical experiment (Fig 2A). All participants exhibited strong alpha peaks in EEG recordings (Fig 2B). With real-time artifact removal, the Kalman filter effectively eliminated stimulation artifacts in EEG recordings (Supp Fig 5). Comparisons with oHT analyses confirmed accurate phase targeting across all participants (Fig 2CD, Supp Fig 6). Both *bias* (F(1.282,5.126) = 0.9724, P = 0.3947, *R*^2^ = 0.1956) and *std* (F(2.246,8.986) = 0.5663, P = 0.6055, *R*^2^ = 0.1240) remained stable across all target phases (Fig 2E). The overall stimulation accuracy across all participants was 81.24% ± 3.53% (Fig 2F) with ±45°tolerance: rising: 78.47% ± 15.48%, peak: 81.66% ± 11.14%, falling: 80.56% ± 12.18%, trough: 81.77% ± 9.78%.

PLDBS accuracy was primarily constrained by system latency (hardware/software jitter plus delay) and oscillation amplitude. Trigger precompensation with pre-triggering signals mitigated fixed 25-ms delays, while residual ∼10-ms jitter limited phase resolution to 4-phase (alpha) and 2-phase (beta) distinctions. Phase estimation errors significantly correlated with oscillation amplitude for each target phase (Fig 3AB, an example from Participant 4, Trough: *P* < 0.0001, *R* = ™0.1993, Rising: *P* < 0.0001, *R* = ™0.2447, peak: *P* < 0.0001, *R* = ™0.3164, Falling: *P* < 0.0001, *R* = ™0.3259).

**Fig 3.**
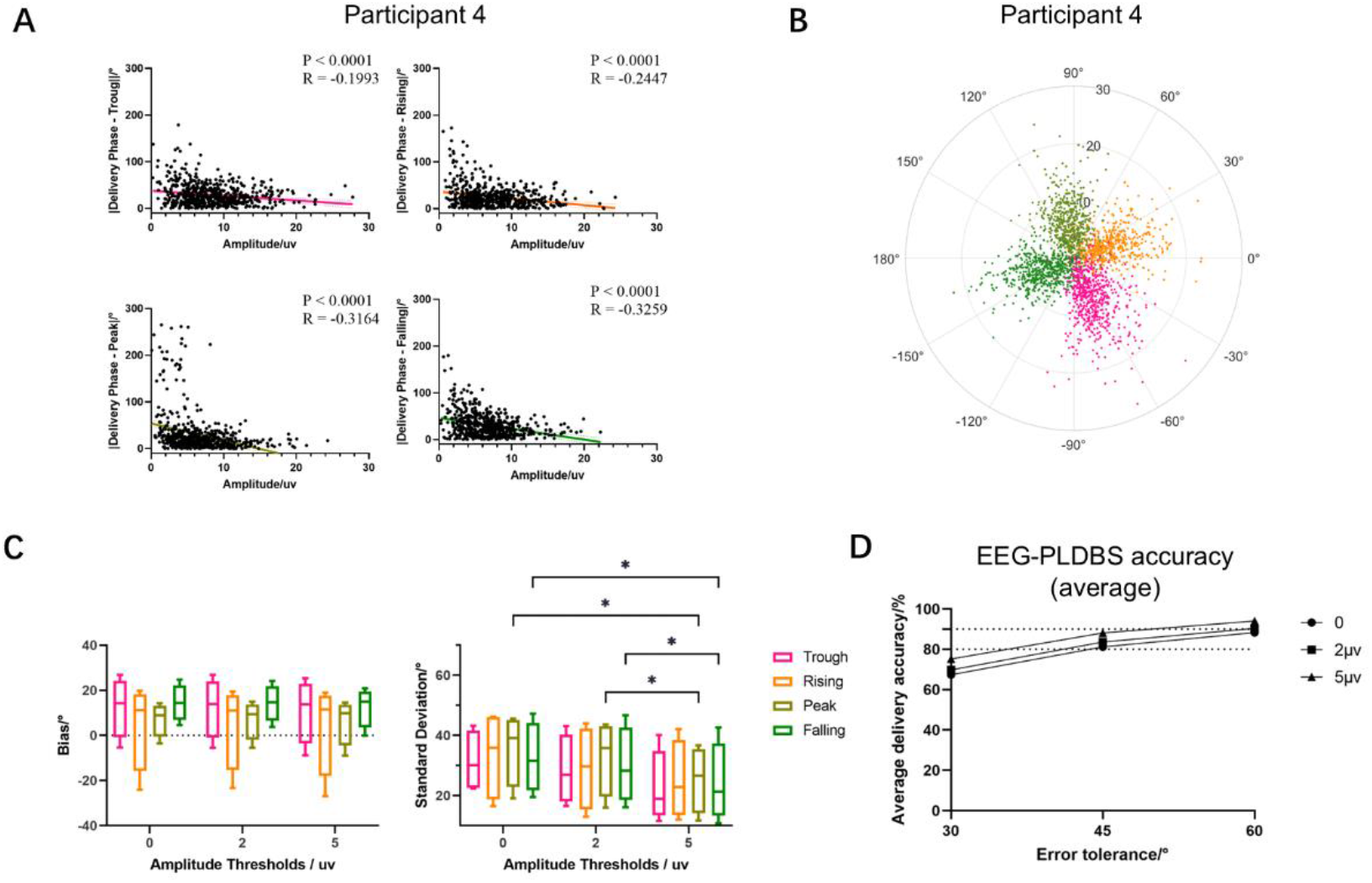
Amplitude-dependent phase targeting optimization. (A) Phase error reduction with amplitude thresholding. Significant negative linear correlations between EEG amplitude and angular deviation were observed across all phases. (B) Relationship between amplitude and delivery phase across all alpha phases in Participant 4. (C) Thresholding efficacy analysis. *Bias* remained consistent when applying amplitude-dependent gating thresholds of 2 *μV* and 5 *μV*, while *std* significantly decreased, particularly in the peak and falling phase conditions. (*: *P* < 0.05). (D) System-wide accuracy improvement. Amplitude gating (5µV threshold) enhanced phase-locking precision from 81.24% to 88.13% (±45°tolerance).

**Fig 4.**
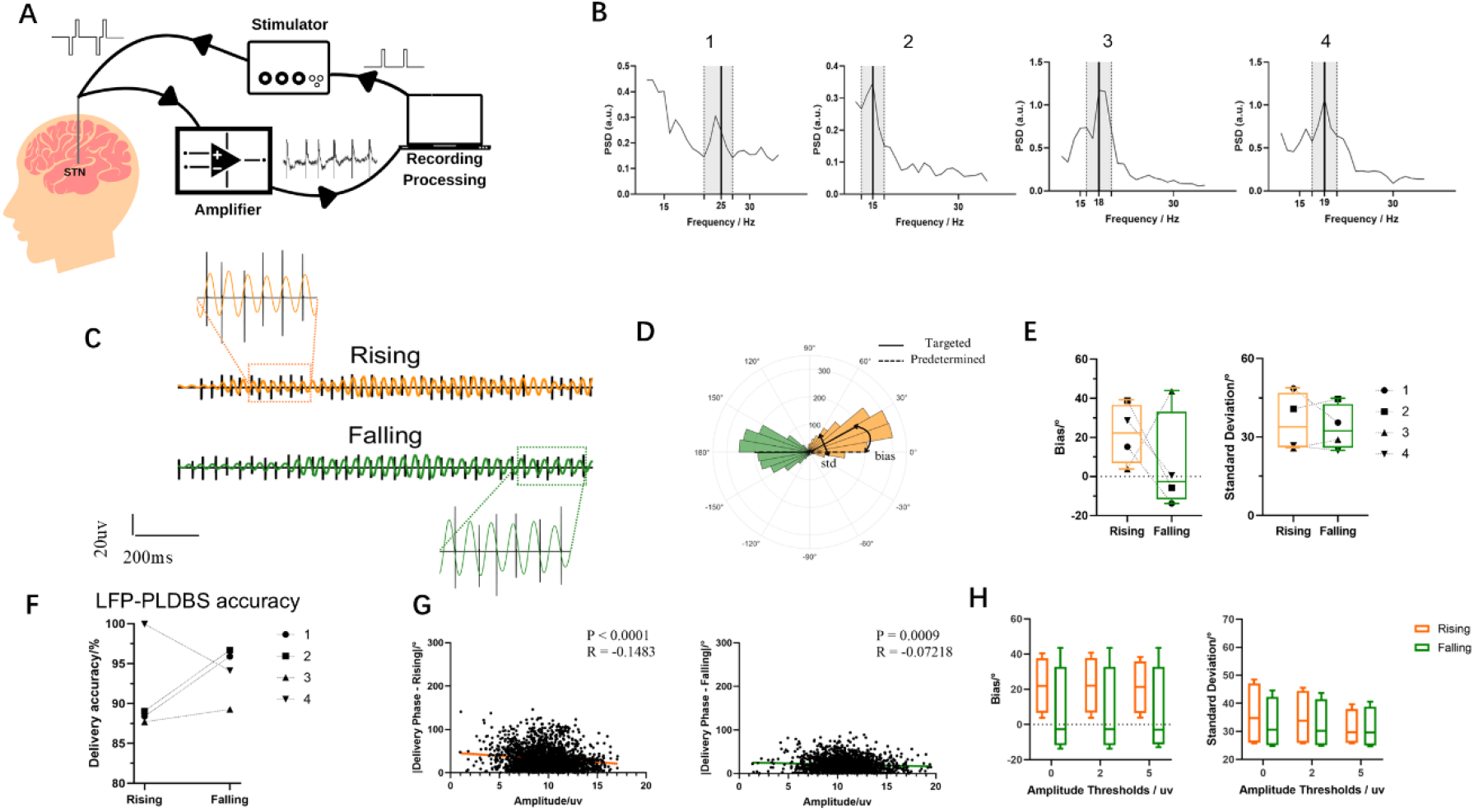
PLDBS of STN beta oscillations. (A) Based on beta oscillations, the PLDBS system uses an LFP channel to deliver electrical pulses to the STN. (B) Individualized beta spectral profiles. Vertical lines indicate patient-specific beta peak frequencies with 5 Hz bandwidth filters (gray-shaded areas). (C) Phase-locked stimulation exemplars (Participant 4). Representative 2-second traces demonstrate quadrant-specific timing at rising (orange) and falling (dark green) phases. Raw LFP with stimulation artifacts are shown in black (left STN). (D) Angular targeting precision. Circular distributions show phase concentration at rising and falling (dashed lines) with mean resultant vectors (black lines) for Participant 4. (E) Cross-participant phase consistency. No significant phase-dependent *bias* have been revealed. (F) LFP-triggered PLDBS demonstrated precise phase-targeting across participants within a ±90°tolerance. (G) Amplitude-error relationship. Significant negative correlations between beta amplitude and phase error. (H) *Bias* and *std* remained consistent when applying amplitude-dependent gating thresholds.

Applying amplitude-dependent gating thresholds of 2 µV and 5 µV to the offline signal resulted in minimal changes in *bias* but a notable reduction in *std* (Fig 3C). A two-way ANOVA confirmed that applying an amplitude threshold had a significant effect on *std* (*F*(1.355,4.065) = 14.46,*P* = 0.0164, *Greenhouse* − *Geisser’s ε* = 0.6775) but not on *bias*(*F*(1.154,3.462) = 1.443, *P* = 0.3157, *Greenhouse* − *Geisser’s ε* = 0.5770), indicating that thresholding reduces variability in phase estimation. Multiple comparison tests demonstrated significant improvements in stimulation *accuracy* for the peak (0 vs. 5 µV (*P* = 0.0207, *t* =) and 2 µV vs. 5 µV (*P* = 0.0158)) and falling (0 vs. 5 µV (*P* = 0.0353) and 2 µV vs. 5 µV (*P* = 0.0247)) phases. With an amplitude threshold of 5 µV, the average stimulation accuracy within a ±45°tolerance across the four targeted phases improved from 81.24% to 88.13% (Fig 3D).

### Real-time STN LFP-triggered PLDBS for precise beta phase targeting

To further explore the clinical potential of the PLDBS system, we conducted experiments using STN beta-triggered PLDBS (Fig 4A). Most stimulation pulses were successfully delivered at the intended target phases (Fig 4CD, Supp Fig 7). *Bias* (*P* = 0.4705, *t* = 0.8237) and *std* (*P* = 0.6501, *t* = 0.5022) remained stable across the two target phases (Fig 4E). The delivery accuracy exceeds 92.64% ± 1.753% (Fig 4F) with ±45°tolernace. Similarly to cortical alpha-triggered PLDBS, estimation error in STN beta-triggered PLDBS significantly correlated with target oscillation amplitude at each phase (Fig 4G, rising: *P* < 0.0001, *R* = ™0.1483, falling: *P* = 0.0009, *R* = ™0.07218). However, when applying amplitude-dependent gating thresholds of 2µV and 5μV, neither *bias* nor *std* showed significant variation (Fig 4H).

### Differential effect of stimulation phase on ERNA

The amplitude of the stimulation-induced ERNA in the STN may indicate the cortico-basal ganglia circuit state^55^ and reflect DBS-mediated engagement of the basal ganglia indirect pathway^54^. Therefore, we analyzed two protocols to explore whether target-phase influences ERNA amplitudes: cortical alpha-triggered PLDBS and STN beta-triggered PLDBS. For cortical alpha-triggered PLDBS (four-phase protocol), a one-way ANOVA was performed within each participant to compare ERNA amplitudes across four targeted phases. No significant phase-specific ERNA amplitude modulation was observed for any participant (Fig 5A; 1: *F* = 0.9556, *P* = 0.4946; 2: *F* = 1.656, *P* = 0.3119; 3: *F* = 0.1521, *P* = 0.9231; 4: *F* = 0.2632, *P* = 0.8492). Similarly, unpaired t-tests comparing ERNA amplitudes between rising and. falling alpha phases (using the STN beta-triggered phase tolerance criteria) showed no significant differences (Fig 5B; 1: *P* = 0.0753, *t* = 3.436; 2: *P* = 0.4496, *t* = 0.9324; 3: *P* = 0.6807, *t* = 0.4675; 4:*P* = 0.5086, *t* = 0.7979). Conversely, STN beta-triggered PLDBS demonstrated significant phase-dependent modulation in 3/4 participants, as assessed by within-subject unpaired t-tests (Fig 5C; 1: *P* = 0.0232, *t* = 3.579; 2: *P* = 0.1454, *t* = 1.085; 3: *P* = 0.0194, *t* = 3.785; 4: *P* = 0.0024, *t* = 6.833). These findings suggest differential recruitment of basal ganglia pathways during phase-targeted stimulation.

**Fig 5.**
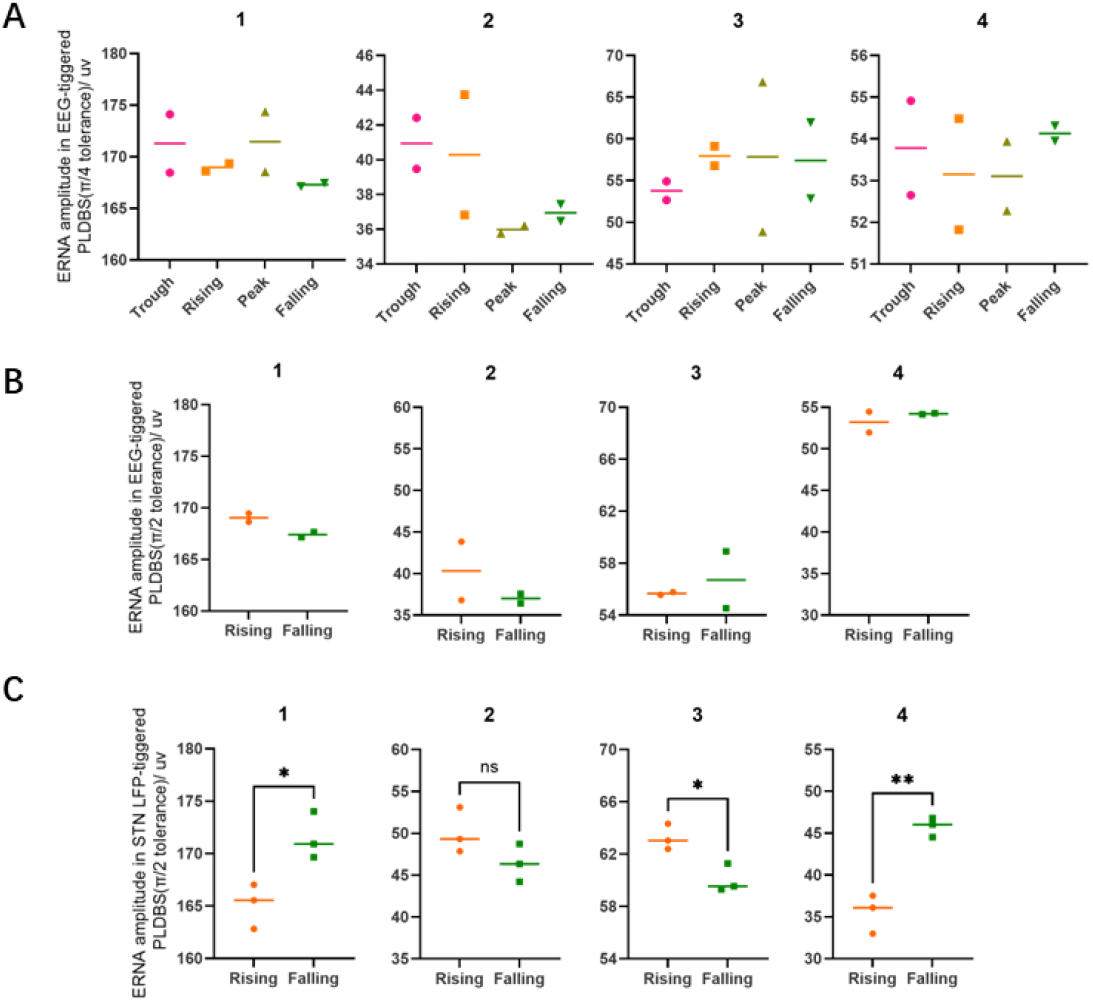
Differential effect of stimulation phase on ERNA amplitudes. (A) No significant phase-locked modulation of ERNA amplitudes was observed across four cortical alpha-phase windows within a ±45°tolerance. (B) Cortical alpha-phase windows (rising vs. falling, ±90°tolerance) revealed no significant differences in ERNA amplitudes between phases. (C) Significant STN beta-phase-dependent modulation emerged in 3/4 participants. (*: *P* < 0.05, **: *P* < 0.01)

### Differential effect of stimulation phase on bradykinesia

Participant 4, who demonstrated the most pronounced ERNA modulation response alongside marked bradykinesia and rigidity symptoms, performed finger-tapping under different stimulation conditions (Fig 6A, Supp Fig 8). Before PLDBS testing, clinical cDBS was applied and significantly increased finger-tapping velocity (Fig 6B, *P* = 0.0009, *t* = 3.399) but not on amplitude (Fig 6C, *P* = 0.2209, *t* = 1.230), though its earlier experimental blocks showed reduced efficacy compared to PLDBS (Fig 6BC). From one-way ANOVA analysis, PLDBS induced phase-specific modulation in both tapping velocity (Fig 6B, *F* = 3.394, *P* = 0.0352) and amplitude (Fig 6C, *F* = 3.252, *P* = 0.043), with rising vs. falling phase differences reaching significance (velocity: *P* = 0.0268; amplitude: *P* = 0.0306). Interestingly, the stimulation phase that elicited greater movement effects also exhibited larger ERNA amplitudes. This phase-locked neuromodulation pattern highlights PLDBS’s optimization potential for symptom-specific targeting.

**Fig 6.**
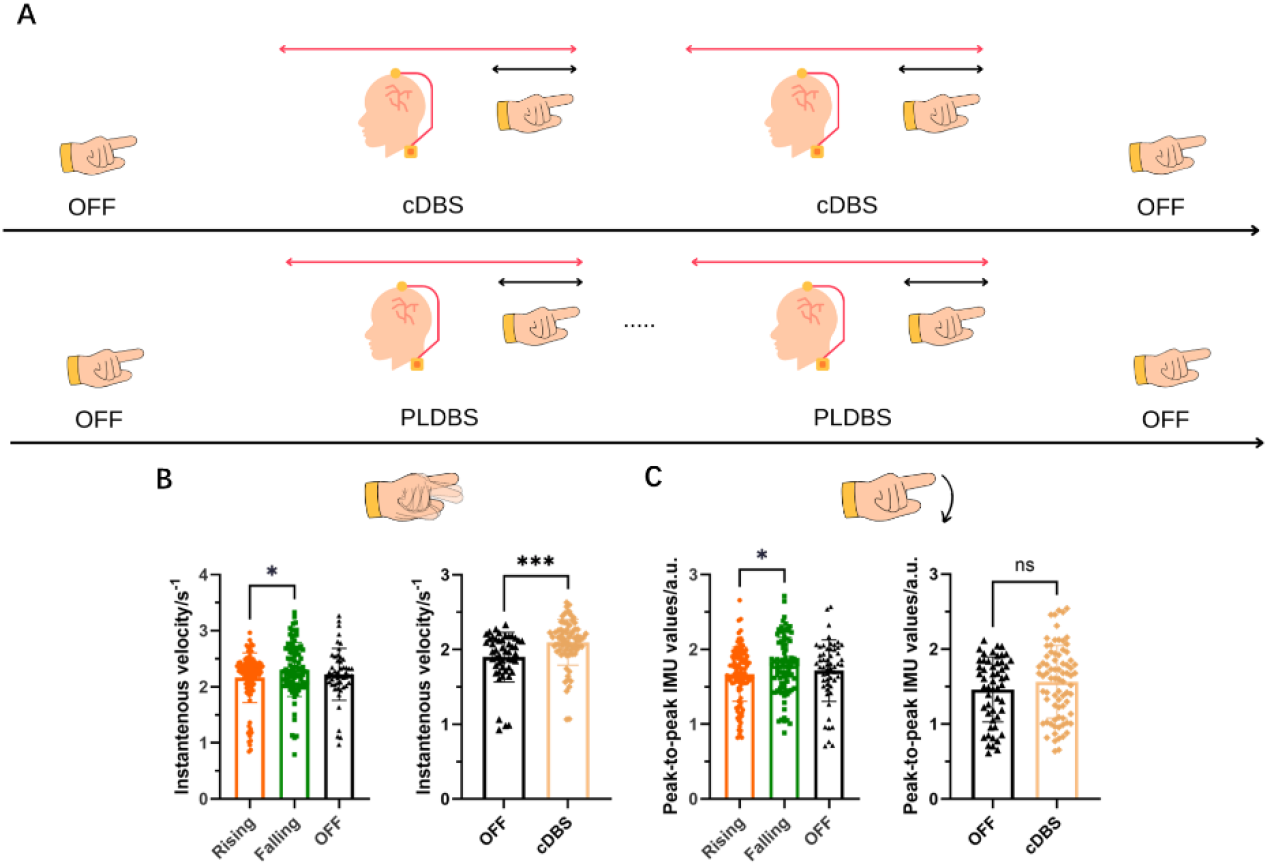
Differential effect of stimulation phase on finger-tapping task performance. (A) Experimental paradigms for cDBS (Top) and PLDBS (Bottom) protocols with finger-tapping tasks. For both paradigms, each block included a 60-second stimulation period (red arrows) followed by a 15-second finger-tapping task (black arrows). In the PLDBS protocol, stimulation pulses were phase-locked to the rising and falling phases of beta oscillations, each for three trials, with phase-targeting order pseudorandomized across trials. Two OFF-stimulation control blocks (pre- and post-intervention) were included to assess baseline effects. (B) Finger-tapping velocity showed differences between the rising and falling phases during PLDBS, and cDBS showed a significant improvement in velocity compared to the OFF-stimulation condition. (C) Finger-tapping amplitudes differed between the rising and falling phases of PLDBS, while no significant difference was found between OFF and cDBS.

## Discussion

In this study, we proposed a PLDBS system that integrates commonly used neurostimulation hardware with a computer-in-the-loop approach. The system was empirically validated as proof-of-concept, demonstrating its potential for flexible and stable stimulation protocols. Our results show that this system can precisely target four distinct phases of cortical alpha oscillations and two distinct phases of STN beta oscillations. Moreover, STN beta-triggered PLDBS revealed phase-dependent effects on ERNA in the STN and motor performance during a finger-tapping task.

### The stability and accuracy of the proposed PLDBS pipeline

The proposed system demonstrates robust phase stimulation capability, enabling precise delivery of four distinct alpha-phase windows (∼10 Hz, 81.24%) and two beta-phase windows (18-25 Hz, 92.64%), with ±45 °tolerance and ±90 °tolerance, respectively. Several factors influence stimulation *accuracy* . First, in Participant 3, tremor activity at 7 Hz broadened stimulation error distributions in EEG-triggered PLDBS (Supp Fig 7) but not in LFP-triggered PLDBS (Supp Fig 8), reducing stimulation accuracy for EEG-based protocols. Therefore, for participants with significant tremors, STN-triggered PLDBS is recommended, while EEG-based approaches require filter design verification to ensure appropriate *bias* and *std* metrics. Second, amplitude thresholding reduces phase error variability in alpha-triggered PLDBS (cortical, 3Hz bandwidth) but not beta-triggered PLDBS (STN, 5Hz bandwidth). Narrower frequency peaks and weaker oscillatory amplitudes in STN beta than cortical alpha oscillations (observed in all 4 cases) may explain this discrepancy. Third, system-induced communication delays create phase estimation *bias*, highlighting the need for latency optimization to bridge simulation-clinical accuracy gaps.

### The flexibility of the PLDBS system

The proposed PLDBS pipeline is flexible and can be easily integrated with different hardware. For instance, it could be effectively implemented using commercially available hardware, such as Neuro Omega systems^24,43^, even without requiring a computer-in-the-loop, instead relying on built-in driver software for real-time recording and stimulation control. Beyond DBS applications, this pipeline could be directly extended to cycle-by-cycle phase-locked transcranial magnetic stimulation (PLTMS), offering an opportunity to investigate the phase-dependent effects of mu-locked PLTMS ^18,20,30,73^. Previous findings suggest a more consistent and shorter intertrial interval could strengthen the relationship between the EEG phase and motor-evoked potential amplitude ^74^. Therefore, cycle-by-cycle PLTMS may offer more insight into this debate.

### Potential movement effects

Our findings revealed ERNA amplitude modulation in STN-PLDBS (Fig 5C), whereas no significant effects were observed in EEG-PLDBS paradigms (Fig 5AB), suggesting STN-PLDBS may reshape neuronal spiking patterns of inhibition^54^. Notably, the observed alignment between ERNA amplitude maxima and peak tapping velocity/amplitude phases supports the importance of the indirect pathway in mediating antiparkinsonian effects^75^. The lack of cortical alpha-mediated modulation could arise from the limited functional significance of the hyper-direct pathway within this specific neuromodulation framework^76^ or the limited selection of targeting phases. Future investigations could prioritize spatiotemporal specificity constraints in phase-domain optimization to elucidate the observed phenomenon, where pre-stimulation biomarker profiling and biophysical modeling^77^ can delineate physiologically optimal intervention phases.

### Limitations

While the PLDBS system demonstrates significant potential, several areas require further refinement. First, the computer-in-the-loop USB architecture introduces inherent jitter and delays, limiting stimulation accuracy. Although our current implementation employed trigger precompensation (advance signal delivery) to mitigate fixed delays, residual jitter (∼10 ms in dedicated stimulation/recording units) persisted as an irreducible constraint. Transitioning to embedded systems with more predictable and stable processing could enhance artifact detection, phase estimation precision, and overall simulation-aligned accuracy. Implementing integrated hardware platforms (e.g., Neuro Omega systems) may optimize temporal precision by minimizing inter-device communication latencies.

Second, while Kalman filtering provides robust stability, the current threshold-based detection method still allows some artifact contamination, which can negatively impact AR model predictions. Enhanced approaches could combine Kalman filtering with real-time interpolation to improve robustness. Additionally, the artifact duration parameter (9 ms) restricts the maximum frequency range that can be effectively targeted, necessitating more flexible algorithms; the SynchroStim artifact removal method could provide a robust approach to mitigating the problems caused by the stimulation pulse artifacts^78^. Further, the low sampling rates of implantable devices^79^ create additional challenges, requiring optimized recording hardware that balances artifact minimization with computational load^78^.

Lastly, the study’s small sample size limits conclusive behavioral impact assessments of PLDBS. While initial results suggest that PLDBS may modulate motor symptoms, its effects were weaker than cDBS, limiting generalizability and highlighting the need for larger clinical trials to confirm PLDBS’s therapeutic potential.

## Conclusions

This study presents a PLDBS system designed for flexible implementation using general neurostimulation devices with a computer-in-the-loop approach. The system effectively targets distinct cortical alpha and STN beta oscillation phases, demonstrating its precision and adaptability. STN beta-triggered stimulation revealed phase-dependent modulation of ERNA amplitudes and finger-tapping performance, suggesting its potential for motor symptom modulation. However, technical refinements are necessary to improve the system further. Transitioning to embedded hardware enabling simultaneous stimulation and faster processing could reduce jitter and delays, improving phase estimation accuracy. These findings underscore PLDBS’s clinical potential as a personalized neuromodulation approach.

## Data Availability

De-identified data from this study will be made accessible on https://data.mrc.ox.ac.uk/mrcbndu/data-sets/search.

## Contributions of authors

Xuanjun Guo: Methodology, Formal analysis, Investigation, Data Curation, Writing—Original Draft, Writing—Review & Editing

Alek Pogosyan: Validation, Investigation, Data Curation

Jean Debarros: Methodology, Formal analysis, Validation, Review & Editing

Shenghong He, Laura Wehmeyer: Validation, Data Curation, Writing—Review & Editing

Fernando Rodriguez Plazas, Ahmed Raslan, Thomas Hart, Francesca Morgante, Keyoumars Ashkan, Erlick A Pereira: Validation, Data Curation

Zixiao Yin: Data Curation, Review & Editing

Karen Wendt: Methodology, Formal analysis

Tim Denison: Methodology, Formal analysis

Andrew Sharrot: Methodology, Investigation

Shouyan Wang: Investigation, Validation, Funding acquisition

Huiling Tan: Methodology, Validation, Investigation, Writing—Original Draft, Writing—Review & Editing, Funding acquisition

## Acknowledgements

This work was supported by the Medical Research Council (MC_UU_00003/2); the Sci-Tech Innovation 2030 Agenda—Major Projects (No. 2021ZD0200407); National Key Research and Development Program of China (No. 2022YFC2405100); the Sci-Tech Innovation 2030 Agenda— Major Projects (No. 2022ZD0205300); National Key Research and Development Program of China (No.2021YFF1200600); National Natural Science Foundation of China (No. 81901153); China Postdoctoral Science Foundation (No. 2019M651373); National Natural Science Foundation of China (No. 82201400); and China Postdoctoral Science Foundation (No. 2022TQ0071). Xuanjun was supported by China Scholarship Council (No. 202306100220). S.H. was supported by the Guarantors of Brain and Royal Society Sino-British Fellowship Trust (IES\R3\213123). We thank all participants for making this study possible.

## Reference

1. Soleimani, G. et al. Closing the loop between brain and electrical stimulation: towards precision neuromodulation treatments. Translational Psychiatry vol. 13 Preprint at 10.1038/s41398-023-02565-5 (2023).

2. Kumari, L. S. & Kouzani, A. Z. Phase-dependent deep brain stimulation: A review. Brain Sciences vol. 11 Preprint at 10.3390/brainsci11040414 (2021).

3. Hussain, S. J. et al. Phase-dependent offline enhancement of human motor memory. Brain Stimul 14, 873–883 (2021).

4. Gersner, R., Kravetz, E., Feil, J., Pell, G. & Zangen, A. Long-term effects of repetitive transcranial magnetic stimulation on markers for neuroplasticity: Differential outcomes in anesthetized and awake animals. Journal of Neuroscience 31, 7521–7526 (2011).

5. Xu, M. et al. Cognitive Effects Following Offline High-Frequency Repetitive Transcranial Magnetic Stimulation (HF-rTMS) in Healthy Populations: A Systematic Review and Meta-Analysis. Neuropsychology Review vol. 34 250–276 Preprint at 10.1007/s11065-023-09580-9 (2024).

6. Pillen, S., Shulga, A., Zrenner, C., Ziemann, U. & Bergmann, T. O. Repetitive sensorimotor mu-alpha phase-targeted afferent stimulation produces no phase-dependent plasticity related changes in somatosensory evoked potentials or sensory thresholds. PLoS One 18, (2023).

7. Jansen, J. M. et al. The Effect of High-Frequency Repetitive Transcranial Magnetic Stimulation on Emotion Processing, Reappraisal, and Craving in Alcohol Use Disorder Patients and Healthy Controls: A Functional Magnetic Resonance Imaging Study. Front Psychiatry 10, (2019).

8. Buzsáki, G. & Draguhn, A. Neuronal Oscillations in Cortical Networks. https://www.science.org (2004).

9. Buzsáki, G., Anastassiou, C. A. & Koch, C. The origin of extracellular fields and currents -EEG, ECoG, LFP and spikes. Nature Reviews Neuroscience vol. 13 407–420 Preprint at 10.1038/nrn3241 (2012).

10. Petersen, P. C. & Buzsáki, G. Cooling of Medial Septum Reveals Theta Phase Lag Coordination of Hippocampal Cell Assemblies. Neuron 107, 731-744.e3 (2020).

11. Peles, O., Werner-Reiss, U., Bergman, H., Israel, Z. & Vaadia, E. Phase-Specific Microstimulation Differentially Modulates Beta Oscillations and Affects Behavior. Cell Rep 30, 2555-2566.e3 (2020).

12. Fiene, M. et al. Phase-specific manipulation of rhythmic brain activity by transcranial alternating current stimulation. Brain Stimul 13, 1254–1262 (2020).

13. Reis, C. et al. Phase-specific Deep Brain Stimulation revisited: effects of stimulation on postural and kinetic tremor. doi:10.1101/2022.06.16.22276451.

14. Santostasi, G. et al. Phase-locked loop for precisely timed acoustic stimulation during sleep. J Neurosci Methods 259, 101–114 (2016).

15. Gordon, P. C., Belardinelli, P., Stenroos, M., Ziemann, U. & Zrenner, C. Prefrontal theta phase-dependent rTMS-induced plasticity of cortical and behavioral responses in human cortex. Brain Stimul 15, 391–402 (2022).

16. Nieuwhof, F. et al. Phase-locked transcranial electrical brain stimulation for tremor suppression in dystonic tremor syndromes. Clinical Neurophysiology 140, 239–250 (2022).

17. Schilberg, L., Oever, S. Ten, Schuhmann, T. & Sack, A. T. Phase and power modulations on the amplitude of TMS-induced motor evoked potentials. PLoS One 16, (2021).

18. Zrenner, C. et al. Corticospinal excitability is highest at the early rising phase of sensorimotor µ-rhythm. Neuroimage 266, (2023).

19. Wischnewski, M., Haigh, Z. J., Shirinpour, S., Alekseichuk, I. & Opitz, A. The phase of sensorimotor mu and beta oscillations has the opposite effect on corticospinal excitability. Brain Stimul 15, 1093–1100 (2022).

20. Torrecillos, F. et al. Motor cortex inputs at the optimum phase of beta cortical oscillations undergo more rapid and less variable corticospinal propagation. Journal of Neuroscience 40, 369–381 (2020).

21. Mansouri, F. et al. A Real-Time Phase-Locking System for Non-Invasive Brain Stimulation. Front Neurosci 12, (2018).

22. Dondé, C. et al. The Effects of Transcranial Electrical Stimulation of the Brain on Sleep: A Systematic Review. Frontiers in Psychiatry vol. 12 Preprint at 10.3389/fpsyt.2021.646569 (2021).

23. Mansouri, F., Dunlop, K., Giacobbe, P., Downar, J. & Zariffa, J. A fast EEG forecasting algorithm for phase-locked transcranial electrical stimulation of the human brain. Front Neurosci 11, (2017).

24. Escobar Sanabria, D. et al. Controlling pallidal oscillations in real-time in Parkinson’s disease using evoked interference deep brain stimulation (eiDBS): Proof of concept in the human. Brain Stimul 15, 1111–1119 (2022).

25. Escobar Sanabria, D. et al. Real-time suppression and amplification of frequency-specific neural activity using stimulation evoked oscillations. Brain Stimul 13, 1732–1742 (2020).

26. Santostasi, G. et al. Phase-locked loop for precisely timed acoustic stimulation during sleep. J Neurosci Methods 259, 101–114 (2016).

27. Ngo, H. V. V., Martinetz, T., Born, J. & Mölle, M. Auditory closed -loop stimulation of the sleep slow oscillation enhances memory. Neuron 78, 545–553 (2013).

28. Neacsiu, A. D. et al. Enhancing Cognitive Restructuring With Concurrent Repetitive Transcranial Magnetic Stimulation: A Transdiagnostic Randomized Controlled Trial. (2021) doi:10.1101/2021.01.18.21250060.

29. Mansouri, F. et al. Effect of Theta Transcranial Alternating Current Stimulation and Phase-Locked Transcranial Pulsed Current Stimulation on Learning and Cognitive Control. Front Neurosci 13, (2019).

30. Bergmann, T. O., Lieb, A., Zrenner, C. & Ziemann, U. Pulsed facilitation of corticospinal excitability by the sensorimotor μ-alpha rhythm. Journal of Neuroscience 39, 10034–10043 (2019).

31. Cagnan, H. et al. Phase dependent modulation of tremor amplitude in essential tremor through thalamic stimulation. Brain 136, 3062–3075 (2013).

32. McNamara, C. G., Rothwell, M. & Sharott, A. Stable, interactive modulation of neuronal oscillations produced through brain-machine equilibrium. Cell Rep 41, (2022).

33. Holt, A. B. et al. Phase-dependent suppression of beta oscillations in parkinson’s disease patients. Journal of Neuroscience 39, 1119–1134 (2019).

34. Miyauchi, E. et al. A novel approach for assessing neuromodulation using phase-locked information measured with TMS-EEG. Sci Rep 9, (2019).

35. Sun, X. et al. Increased Entrainment and Decreased Excitability Predict Efficacious Treatment of Closed-Loop Phase-Locked RTMS for Treatment-Resistant Depression.

36. Peles, O., Werner-Reiss, U., Bergman, H., Israel, Z. & Vaadia, E. Phase-Specific Microstimulation Differentially Modulates Beta Oscillations and Affects Behavior. Cell Rep 30, 2555-2566.e3 (2020).

37. Lio, G., Thobois, S., Ballanger, B., Lau, B. & Boulinguez, P. Removing deep brain stimulation artifacts from the electroencephalogram: Issues, recommendations and an open-source toolbox. Clinical Neurophysiology vol. 129 2170–2185 Preprint at 10.1016/j.clinph.2018.07.023 (2018).

38. Swann, N. C. et al. Adaptive deep brain stimulation for Parkinson’s disease using motor cortex sensing. J Neural Eng 15, (2018).

39. Quinn, E. J. et al. Beta oscillations in freely moving Parkinson’s subjects are attenuated during deep brain stimulation. Movement Disorders 30, 1750–1758 (2015).

40. Neumann, W. J., Gilron, R., Little, S. & Tinkhauser, G. Adaptive Deep Brain Stimulation: From Experimental Evidence Toward Practical Implementation. Movement Disorders vol. 38 937–948 Preprint at 10.1002/mds.29415 (2023).

41. Qian, X. et al. A method for removal of deep brain stimulation artifact from local field potentials. IEEE Transactions on Neural Systems and Rehabilitation Engineering 25, 2217– 2226 (2017).

42. Heffer, L. F. & Fallon, J. B. A novel stimulus artifact removal technique for high-rate electrical stimulation. J Neurosci Methods 170, 277–284 (2008).

43. Nie, Y. et al. Real-time removal of stimulation artifacts in closed-loop deep brain stimulation. J Neural Eng 18, (2021).

44. Zhou, A., Johnson, B. C. & Muller, R. Toward true closed-loop neuromodulation: artifact-free recording during stimulation. Current Opinion in Neurobiology vol. 50 119–127 Preprint at 10.1016/j.conb.2018.01.012 (2018).

45. Boashash, B. Estimating and Interpreting the Instantaneous Frequency of a Signal-Part 2: Algorithms and Applications. (1992).

46. Zanos, S., Rembado, I., Chen, D. & Fetz, E. E. Phase-Locked Stimulation During Cortical Beta Oscillations Produces Bidirectional Synaptic Plasticity in Awake Monkeys. Current Biology 28, 2515-2526.e4 (2018).

47. Rosenblum, M. G. et al. Locking-Based Frequency Measurement and Synchronization of Chaotic Oscillators with Complex Dynamics. Phys Rev Lett 89, (2002).

48. Zrenner, C. et al. The shaky ground truth of real-time phase estimation. Neuroimage 214, (2020).

49. Schreglmann, S. R. et al. Non-invasive suppression of essential tremor via phase-locked disruption of its temporal coherence. Nat Commun 12, (2021).

50. Wodeyar, A., Schatza, M., Widge, A. S., Eden, U. T. & Kramer, M. A. A state space modeling approach to real-time phase estimation. (2021) doi:10.7554/eLife.

51. Wodeyar, A., Marshall, F. A., Chu, C. J., Eden, U. T. & Kramer, M. A. Different Methods to Estimate the Phase of Neural Rhythms Agree But Only During Times of Low Uncertainty. eNeuro 10, (2023).

52. Rosenblum, M., Pikovsky, A., Kühn, A. A. & Busch, J. L. Real -time estimation of phase and amplitude with application to neural data. Sci Rep 11, (2021).

53. Wang, S. et al. Closed-Loop Adaptive Deep Brain Stimulation in Parkinson’s Disease: Procedures to Achieve It and Future Perspectives. Journal of Parkinson’s Disease vol. 13 453–471 Preprint at 10.3233/JPD-225053 (2023).

54. Steiner, L. A. et al. Neural signatures of indirect pathway activity during subthalamic stimulation in Parkinson’s disease. Nat Commun 15, (2024).

55. Wiest, C. et al. Evoked resonant neural activity in subthalamic local field potentials reflects basal ganglia network dynamics. Neurobiol Dis 178, (2023).

56. Wiest, C. et al. Local field potential activity dynamics in response to deep brain stimulation of the subthalamic nucleus in Parkinson’s disease. Neurobiol Dis 143, (2020).

57. 42nd Annual International Conferences of the IEEE Engineering in Medicine and Biology Society : ‘Enabling Innovative Technologies for Global Healthcare’ : 20-24 July 2020, Montreal, Canada. (IEEE, 2020).

58. Morbidi, F., Garulli, A., Prattichizzo, D., Rizzo, C. & Rossi, S. Application of Kalman filter to remove TMS-induced artifacts from EEG recordings. IEEE Transactions on Control Systems Technology 16, 1360–1366 (2008).

59. Morbidi, F. et al. Off-line removal of TMS-induced artifacts on human electroencephalography by Kalman filter. J Neurosci Methods 162, 293–302 (2007).

60. Hussain, S. J. et al. Phase-dependent offline enhancement of human motor memory. Brain Stimul 14, 873–883 (2021).

61. Schaworonkow, N., Triesch, J., Ziemann, U. & Zrenner, C. EEG-triggered TMS reveals stronger brain state-dependent modulation of motor evoked potentials at weaker stimulation intensities. Brain Stimul 12, 110–118 (2019).

62. Busch, J. L., Feldmann, L. K., Kühn, A. A. & Rosenblum, M. Real -time phase and amplitude estimation of neurophysiological signals exploiting a non-resonant oscillator. Exp Neurol 347, (2022).

63. Wiest, C. et al. Local field potential activity dynamics in response to deep brain stimulation of the subthalamic nucleus in Parkinson’s disease. Neurobiol Dis 143, (2020).

64. Rosenblum, M., Pikovsky, A., Kühn, A. A. & Busch, J. L. Real -time estimation of phase and amplitude with application to neural data. Sci Rep 11, (2021).

65. Thies, M., Zrenner, C., Ziemann, U. & Bergmann, T. O. Sensorimotor mu-alpha power is positively related to corticospinal excitability. Brain Stimul 11, 1119–1122 (2018).

66. Bergmann, T. O., Lieb, A., Zrenner, C. & Ziemann, U. Pulsed facilitation of corticospinal excitability by the sensorimotor μ-alpha rhythm. Journal of Neuroscience 39, 10034–10043 (2019).

67. Little, S. & Brown, P. The functional role of beta oscillations in Parkinson’s disease. Parkinsonism Relat Disord 20 Suppl 1, (2014).

68. Reis, C. et al. Phase-specific Deep Brain Stimulation revisited: effects of stimulation on postural and kinetic tremor. Preprint at 10.1101/2022.06.16.22276451 (2022).

69. Fischer, P. et al. Entraining stepping movements of Parkinson’s patients to alternating subthalamic nucleus deep brain stimulation. Journal of Neuroscience 40, 8964–8972 (2020).

70. McNamara, C. G., Rothwell, M. & Sharott, A. Stable, interactive modulation of neuronal oscillations produced through brain-machine equilibrium. Cell Rep 41, (2022).

71. Salimpour, Y., Mills, K. A., Hwang, B. Y. & Anderson, W. S. Phase-Targeted Stimulation Modulates Phase-Amplitude Coupling in the Motor Cortex of the Human Brain. Brain Stimul 15, 152–163 (2022).

72. Wiest, C. et al. Subthalamic Nucleus Stimulation–Induced Local Field Potential Changes in Dystonia. Movement Disorders 38, 423–434 (2023).

73. Desideri, D., Zrenner, C., Ziemann, U. & Belardinelli, P. Phase of sensorimotor μ-oscillation modulates cortical responses to transcranial magnetic stimulation of the human motor cortex. Journal of Physiology 597, 5671–5686 (2019).

74. Madsen, K. H. et al. No trace of phase: Corticomotor excitability is not tuned by phase of pericentral mu-rhythm. Brain Stimul 12, 1261–1270 (2019).

75. Mastro, K. J. et al. Cell-specific pallidal intervention induces long-lasting motor recovery in dopamine-depleted mice. Nat Neurosci 20, 815–823 (2017).

76. Johnson, L. A. et al. Direct activation of primary motor cortex during subthalamic but not pallidal deep brain stimulation. Journal of Neuroscience 40, 2166–2177 (2020).

77. Duchet, B. & Bogacz, R. How to design optimal brain stimulation to modulate phase-amplitude coupling? J Neural Eng 21, (2024).

78. Jean Debarros; Lea Gaignon; Shenghong He; Alek Pogosyan; Moaad Benjaber; Timothy Denison. Artefact-free recording of local field potentials with simultaneous stimulation for closed-loop Deep-Brain Stimulation. (2020) doi:10.1109/EMBC44109.2020.9176665.

79. Stephen L. Schmidt, 1,† Afsana H. Chowdhury,2,† Kyle T. Mitchell,3,† Jennifer J. Peters,1 Qitong Gao,2 Hui-Jie Lee,4 Katherine Genty,5 Shein-Chung Chow,4 Warren M. Grill,1,2,5,6 Miroslav Pajic2 and Dennis A. Turner1,5,6. At home adaptive dual target deep brain stimulation in Parkinson’s disease with proportional control. Brain 147, 749–751 (2024).

