## Supplementary Table 1 - 3, Figure 1 - 8 for "Flexible and Stable Cycle-by-Cycle Phase-Locked Deep Brain Stimulation System Targeting Brain Oscillations in the Management of Movement Disorders"

**Real-time artifact removal method**

Bandpass filter is a simple approach to remove artifacts. In power-based adaptive DBS scenarios^1^, an IIR filter is often used to isolate beta oscillations during high-frequency DBS ^2–4^. However, this method is unsuitable for phase-locked stimulation because the targeted frequency range overlaps with the frequency of the stimulation pulses. Template subtraction methods have gained significant interest and are widely applied in phase-locked clinical studies ^5–8^. However, these methods assume that stimulation artifacts have a constant pattern, which is often not the case in practice due to variability in stimulation pulse characteristics. An alternative approach is irregular sampling^9^ or “blanking”, where recorded data during the stimulation artifact interval is removed and replaced. While effective in real-time clinical recordings, this method introduces discontinuity in the signal and down-sampling can result in data loss, especially with prolonged stimulation artifacts. Moreover, it is highly sensitive to errors in detecting artifact onset, particularly in systems with processing delays.

**Artifact removal method validation**

We evaluated the stability and efficacy of the Kalman filter and irregular sampling methods through simulations using a generated signal containing artifacts with pre-determined artifacts durations and variations. The signal was generated as described in a previous paper^9^. The original recording consisted of STN local filed potentials (LFPs) from a PD patient with significant beta activity (peak frequency: 20-24Hz) in the OFF-medication and OFF-stimulation condition. Stimulation artifacts were simulated by post-fusing the LFP recordings containing real DBS artifacts recorded from the same patient. Specifically, 12 ms of recordings following 25 individual stimulation pulses were extracted and added to the original signal each time a specific phase was detected. The simulation framework mimics cycle-by-cycle PLDBS dynamics, closely replicating clinical trial conditions.

To assess the effectiveness of the irregular sampling and Kalman filter methods, we measured the relative error between the recovered signals and original recordings. The power spectral density (PSD) of both signals was estimated using Welch's method with a 1-second time window. The relative error in peak frequency band was calculated across various pre-determined artifacts durations, following the approach described in a previous paper ^9^ :

$$relative error= \frac{1}{N}\sum\frac{\left| p_{f}\left( f \right)-p_{t}(f) \right|}{\left| p_{t}(f) \right|}\times100\%$$

**Real-time phase estimation method**

One class of phase estimation methods can be broadly categorized into direct phase estimation techniques that do not require analytic signal reconstruction and estimate only the phase and not the envelope of the signal. One widely used approach is the zero-crossing estimation method ^10,11^, which determines the instantaneous cycle frequency by measuring intervals between zero-crossings. The phase of the current time point is then calculated based on the time difference between the current point and previous zero-crossing point. Another approach involves phase-lock oscillators^12^, which can be further enhanced by applying a low-pass filter to form a Phase-Locking Loop (PLL) ^13^. Both phase-lock oscillators and PLL rely on phase errors between a reference and input signal to dynamically adjust their parameters for improved phase estimation.

In recent years, a second class of phase estimation methods has increasingly focused on deriving analytic signals from the real part of the signal. Estimation errors are assessed by comparing the real part of the estimated analytic signal with the actual signal. One widely used approach is the HT, which extracts both phase and envelope after applying an FIR filter ^8^. However, since HT is inherently non-causal, making it unsuitable for real-time applications, alternative causal techniques have been developed, such as the endpoint-corrected HT ^14^ (ecHT) and autoregressive HT (arHT) ^15–18^.

The third class of phase estimation methods employs more complex mathematical models and algorithms to = reconstruct analytic signals. Such approaches integrate state-space models or resonance theory. State-space models predict the future data points using state equations, dynamically update phase parameters by rotating the current state and adjusting estimates based on the discrepancies between predicted and actual values. Examples include the state space phase estimator (SSPE) ^19^ and the simplified state-space model OscillTrack ^20^. Based on resonance theory, resonant oscillators (RO) and non-resonant oscillators (NRO) utilize two constructed oscillators, one tracking the input amplitude and the other matching to the input phase ^13,21^. NRO employs a linear damped oscillator for phase estimation, while RO relies on a linear oscillator. Compared to RO, NRO covers a broader frequency range and achieves higher estimation accuracy in neural oscillation signals.

| Publication | Methodology | Application |
| --- | --- | --- |
| Little et al., 2013 | Bandpass filter | Swann et al., 2018, Quinn et al.,2015, Neumann et al., 2023 |
| Morbidi et al., 2007 | Kalman filter | Morbidi et al., 2008 |
| Hashimotoet al., 2001, Sun and Hinrichs, 2016, Qian et al., 2017, Liu et al., 2024 | Template subtraction | Chen et al., 2019, Sanabria et al., 2020, Sanabria et al., 2022 |
| Heffer and Fallon, 2008, Nie et al., 2021 | Interpolation or sampling | Zhou et al., 2018, McNamara et al., 2022 |

Table 1 Review of artifacts removal methodology

Table 2 Review of real-time phase estimation methodology

| Publication | Methodology |  |
| --- | --- | --- |
| Boashash et al., 1992, Zanos et al., 2018 | **Zero-crossing** | $\boldsymbol{\varphi=}\frac{\boldsymbol{t -}\boldsymbol{t}_{\boldsymbol{last\_ZC}}}{\boldsymbol{T}_{\boldsymbol{ZC}}}$ |
| Rosenblum et al., 2002, Santostasi et al., 2016 | Phase-lock oscillators or phase locking loop | $\dot{\boldsymbol{\theta}}\boldsymbol{=\omega-\varepsilon sin\theta\cdot s(t)}$;$\boldsymbol{\theta\approx\varphi}$ |
| Montaseri et al., 2013 | Resonant oscillators | $\tilde{\boldsymbol{x}}\boldsymbol{(t)=}\ddot{\boldsymbol{s}}\boldsymbol{+\alpha}\dot{\boldsymbol{s}}\boldsymbol{+}\boldsymbol{\omega}^{\boldsymbol{2}}\boldsymbol{s}$; $\dot{\boldsymbol{s}}\boldsymbol{=\mu}\tilde{\boldsymbol{x}}\left( \boldsymbol{t} \right)\boldsymbol{+}\tilde{\boldsymbol{x}}\boldsymbol{(t)}$ |
| Mansouri et al., 2017, Zrenner et al., 2020, Schreglmann et al., 2021, Takeuchi et al., 2021 | Hilbert Transform and Casual Hilbert Transform (e.g. **arHT**, ecHT) | $\tilde{\boldsymbol{x}}\boldsymbol{(t)=x}\left( \boldsymbol{t} \right)\boldsymbol{+j}\boldsymbol{\cdot}\boldsymbol{x}_{\boldsymbol{Hilbert}}\boldsymbol{(t)}$ |
| Anirudh et al., 2021 | **State space phase estimator (SSPE)** | $\tilde{\boldsymbol{x}}\left( \boldsymbol{t} \right)\boldsymbol{=a\theta}\tilde{\boldsymbol{x}}\left( \boldsymbol{t-1} \right) \boldsymbol{+\mu(t)}$ |
| Rosenblum et al., 2021 | **Non resonant oscillators (NRO)** | $\tilde{\boldsymbol{x}}\boldsymbol{(t)=}\ddot{\boldsymbol{s}}\boldsymbol{+\alpha}\dot{\boldsymbol{s}}\boldsymbol{+}\boldsymbol{\omega}^{\boldsymbol{2}}\boldsymbol{s}$ |
| McNamara et al., 2022 | **OscillTrack** | $\tilde{\boldsymbol{x}}\left( \boldsymbol{t} \right)\boldsymbol{=}\boldsymbol{\alpha}_{\boldsymbol{n}}\boldsymbol{sin}\boldsymbol{\omega}_{\boldsymbol{t}}\boldsymbol{+}\boldsymbol{\beta}_{\boldsymbol{n}}\boldsymbol{cos}\boldsymbol{\omega}_{\boldsymbol{t}}\boldsymbol{+j(}\boldsymbol{\beta}_{\boldsymbol{n}}\boldsymbol{sin}\boldsymbol{\omega}_{\boldsymbol{t}}\boldsymbol{-}\boldsymbol{\alpha}_{\boldsymbol{n}}\boldsymbol{cos}\boldsymbol{\omega}_{\boldsymbol{t}}\boldsymbol{)}$ |

Table 3 Quantification of artifacts removal

| Relative errors | 8ms | 9ms | 10ms | 11ms | 12ms |
| --- | --- | --- | --- | --- | --- |
| Irregular sampling | 409.91% | 14.56% | 13.52% | 11.41% | 10.84% |
| Kalman filter | **36.18%** | **5.21%** | **5.56%** | **5.62%** | **5.84%** |

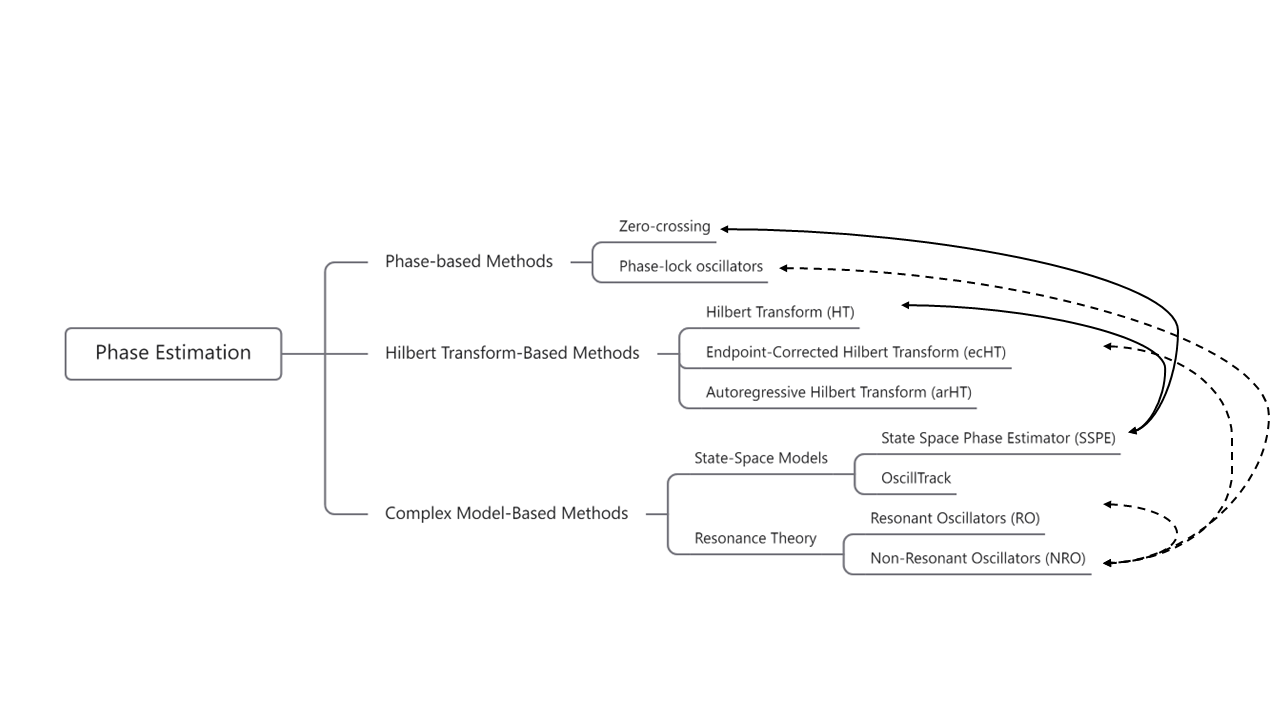

Figure 1 Several studies compared current real-time phase estimation methods.

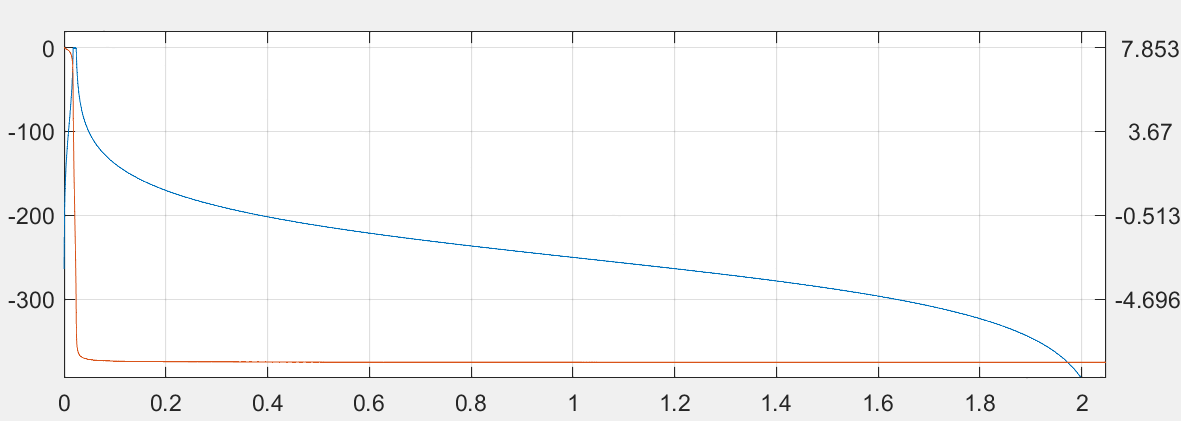

Frequency/ $\times\pi$ rad/sample

Phase /rad

Magnitude /dB

Figure 2 Frequency response characteristics of a fourth-order Chebyshev IIR filter.

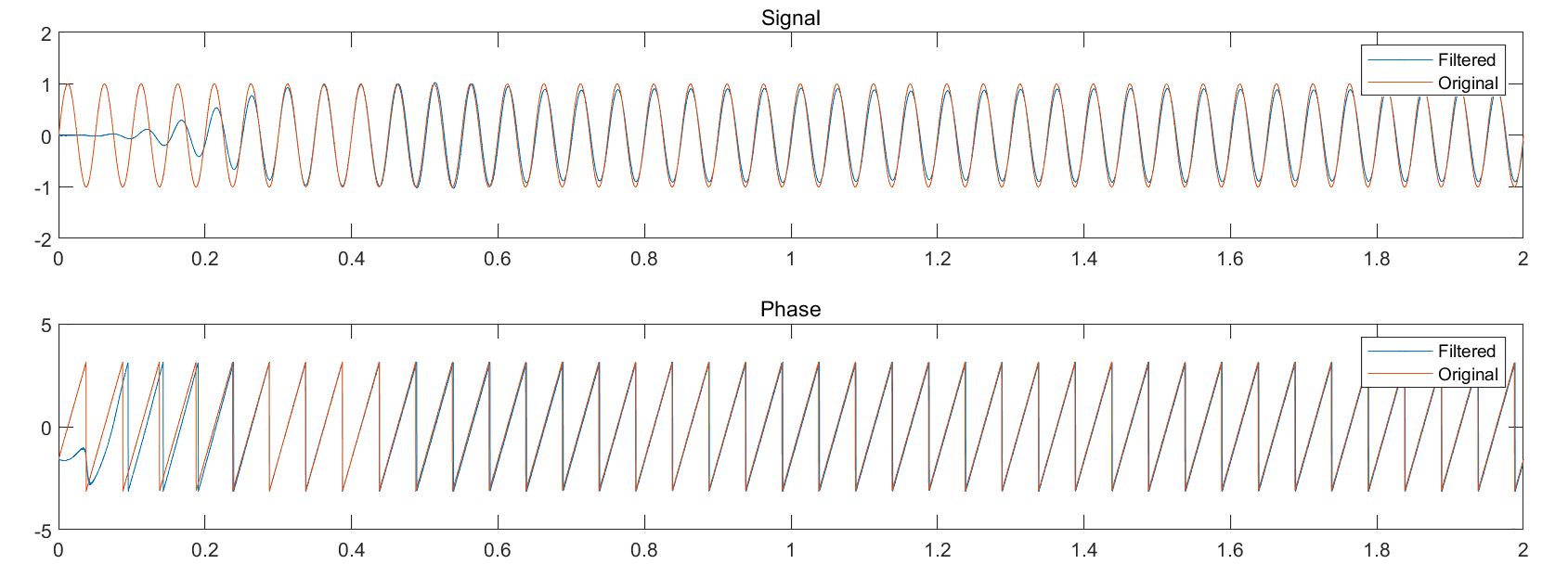

Figure 3 The sine wave processed by the IIR filter retains most of its amplitude information (Up) and exhibits an approximately linear phase response after shifting (Down).

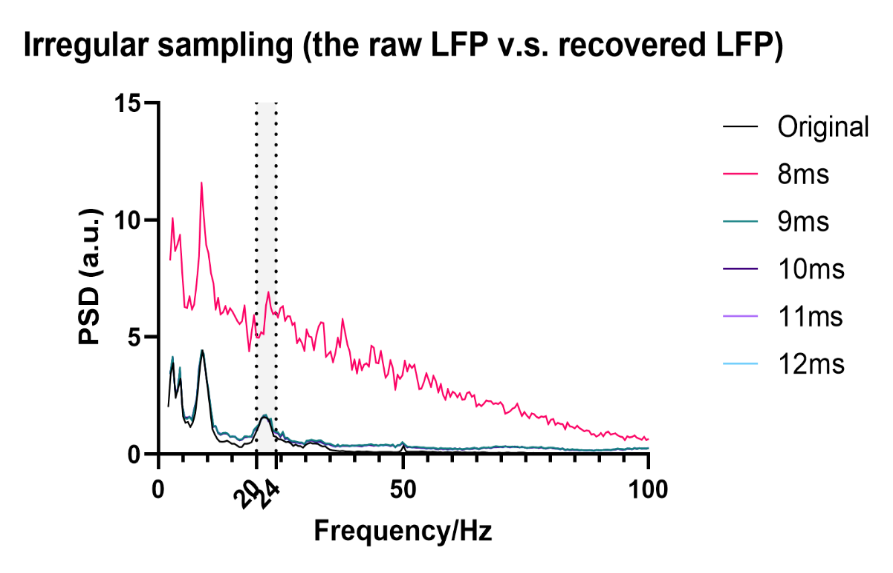

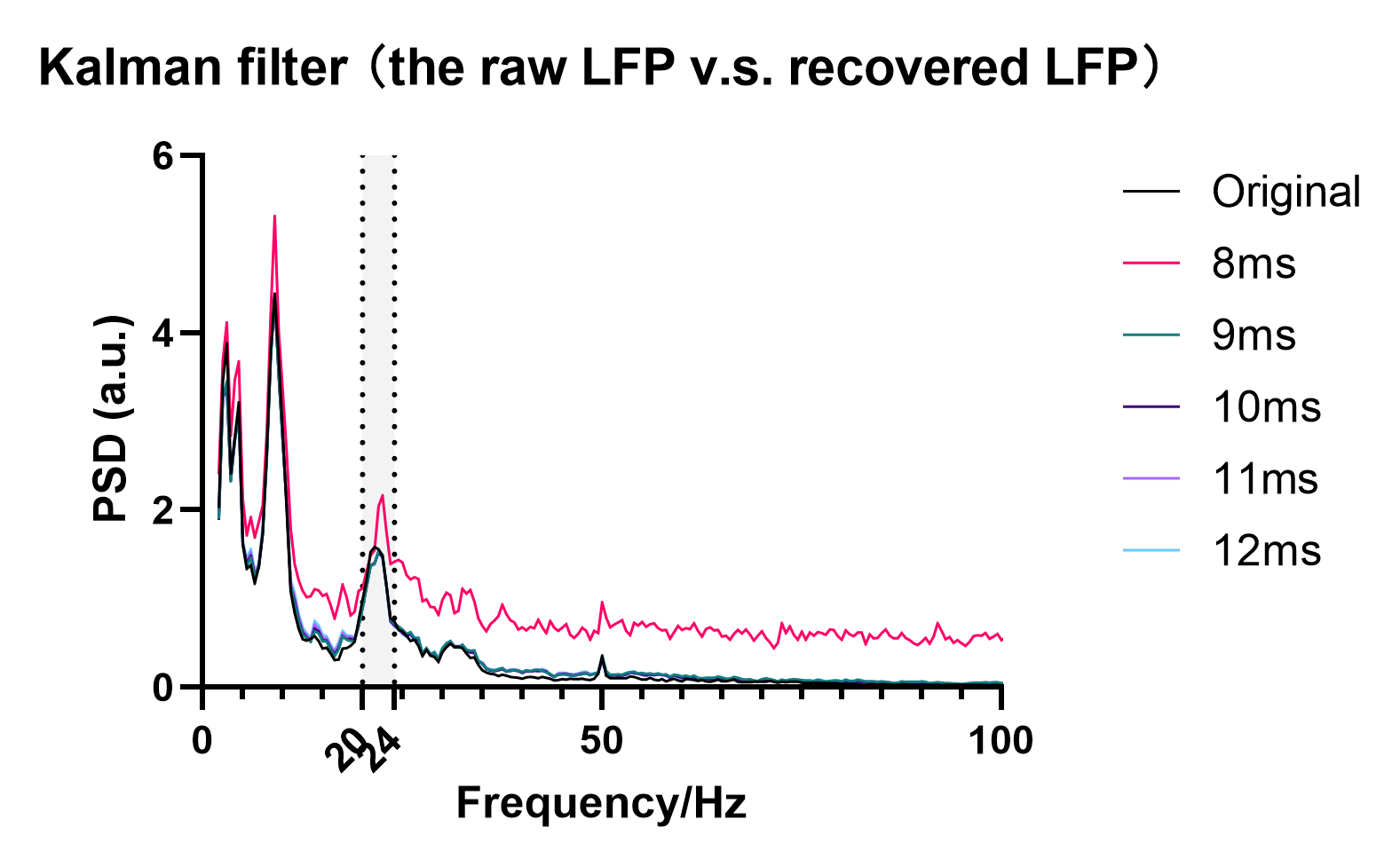

Figure 4 Comparative power spectral density analysis of contaminated signals under irregular sampling method versus Kalman-filtered method across varying stimulation durations.

(Top) Irregular sampling results; (Bottom) Kalman-filter processed signals.

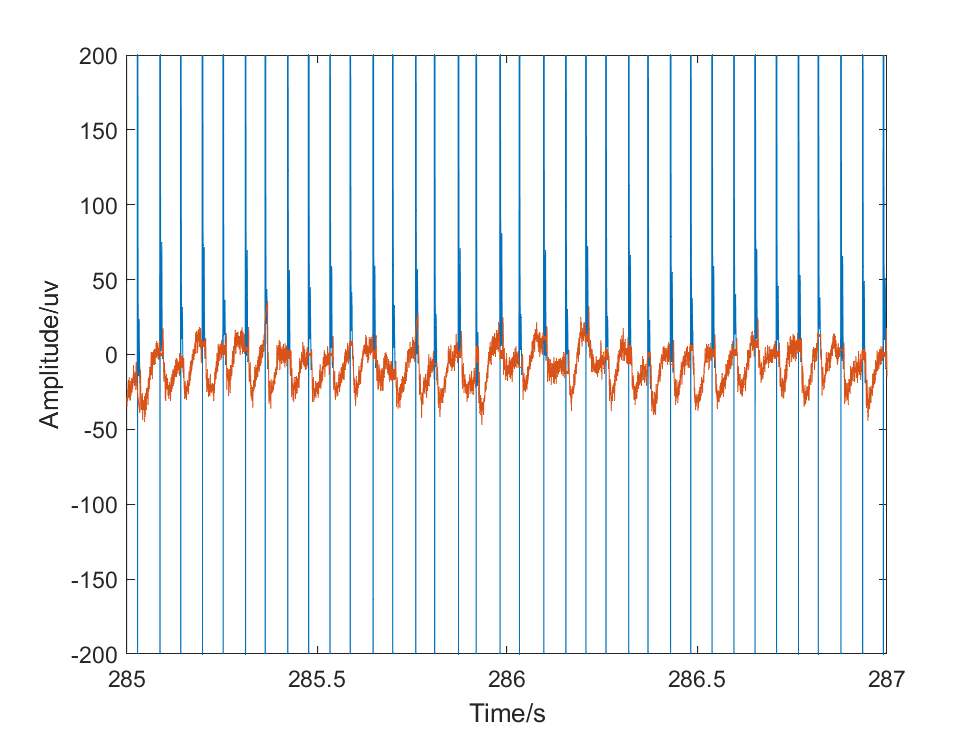

Figure 5 Real-Time Artifact Removal Process. Comparison of the signal processed by the Kalman filter (orange) and the raw signal (blue). The Kalman filter effectively mitigates artifacts, showcasing its role in improving signal quality for real-time phase estimation.

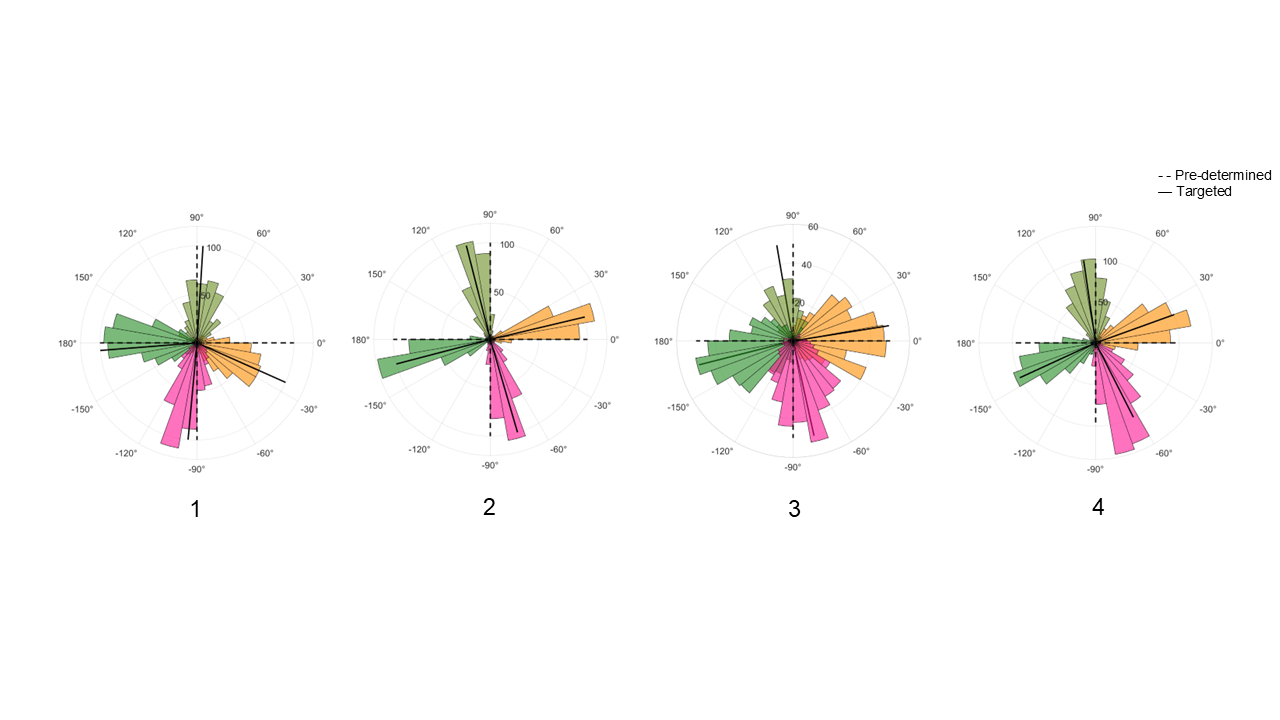

Figure 6 Delivery phase distribution in all participants on cortical alpha PLDBS.

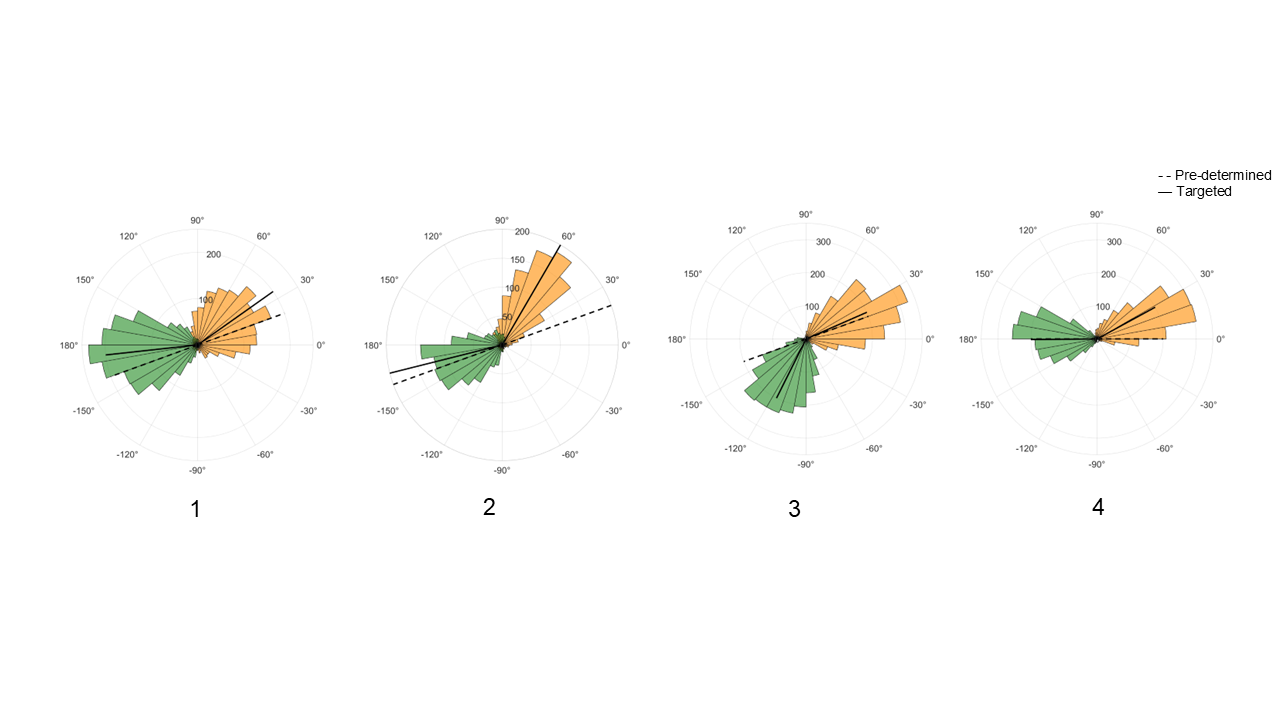

Figure 7 Delivery phase distribution in all participants on STN beta PLDBS.

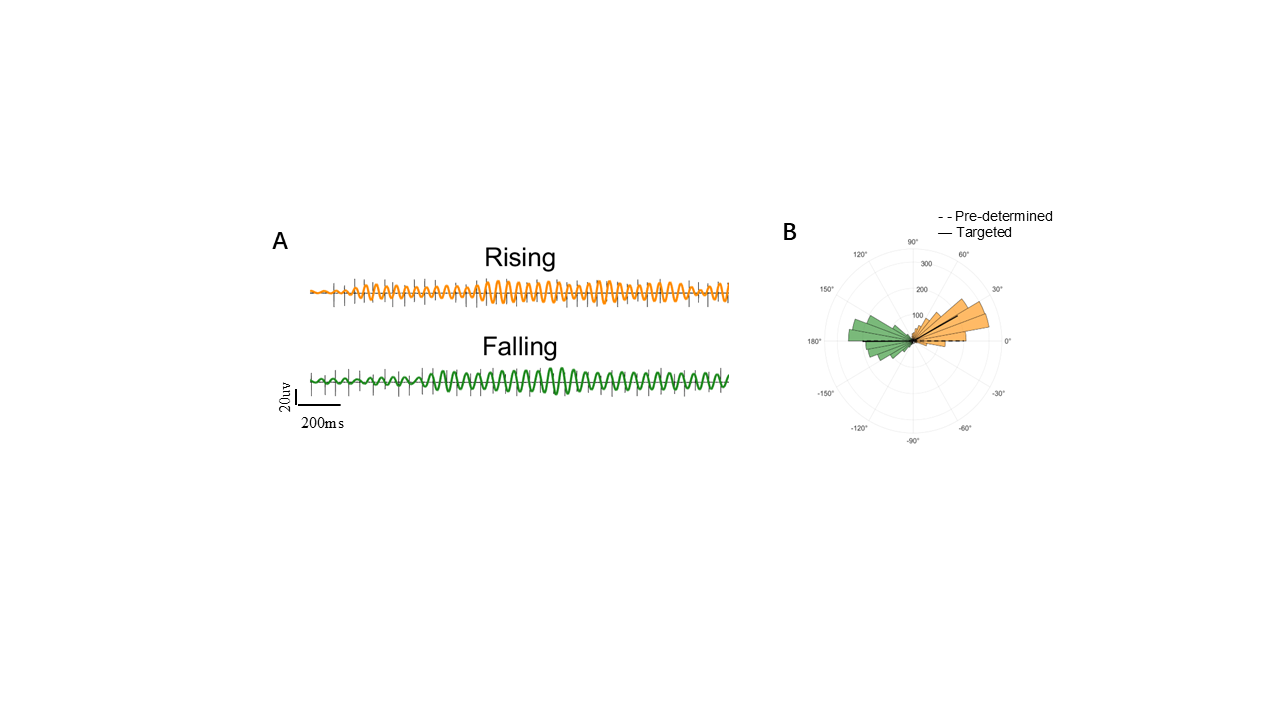

Figure 8 Stimulation and rose plot of finger-tapping task (A) Example of 2 s signals across two phase conditions during finger tapping tasks. Pulses are delivered at the rising (orange and falling (dark green) beta phases. Black lines represent raw wideband signals which contain stimulation artifacts. (B) Distribution of delivery phases during tapping tasks across two targeted phases. The black lines represent the mean values of the distributions and dashed lines indicate the pre-determined phase.

**Reference**

1.	Little, S. *et al.* Adaptive deep brain stimulation in advanced Parkinson disease. *Ann Neurol* 74, 449–457 (2013).

2.	Quinn, E. J. *et al.* Beta oscillations in freely moving Parkinson’s subjects are attenuated during deep brain stimulation. *Movement Disorders* 30, 1750–1758 (2015).

3.	Swann, N. C. *et al.* Adaptive deep brain stimulation for Parkinson’s disease using motor cortex sensing. *J Neural Eng* 15, (2018).

4.	Neumann, W. J., Gilron, R., Little, S. & Tinkhauser, G. Adaptive Deep Brain Stimulation: From Experimental Evidence Toward Practical Implementation. *Movement Disorders* vol. 38 937–948 Preprint at https://doi.org/10.1002/mds.29415 (2023).

5.	Qian, X. *et al.* A method for removal of deep brain stimulation artifact from local field potentials. *IEEE Transactions on Neural Systems and Rehabilitation Engineering* 25, 2217–2226 (2017).

6.	Yuan, H. *et al.* Detection and Quantification of Resting Tremor in Parkinson’s Disease Using Long-Term Acceleration Data. *Math Probl Eng* 2021, (2021).

7.	Escobar Sanabria, D. *et al.* Real-time suppression and amplification of frequency-specific neural activity using stimulation evoked oscillations. *Brain Stimul* 13, 1732–1742 (2020).

8.	Escobar Sanabria, D. *et al.* Controlling pallidal oscillations in real-time in Parkinson’s disease using evoked interference deep brain stimulation (eiDBS): Proof of concept in the human. *Brain Stimul* 15, 1111–1119 (2022).

9.	Nie, Y. *et al.* Real-time removal of stimulation artifacts in closed-loop deep brain stimulation. *J Neural Eng* 18, (2021).

10.	Zanos, S., Rembado, I., Chen, D. & Fetz, E. E. Phase-Locked Stimulation During Cortical Beta Oscillations Produces Bidirectional Synaptic Plasticity in Awake Monkeys. *Current Biology* 28, 2515-2526.e4 (2018).

11.	Boashash, B. *Estimating and Interpreting the Instantaneous Frequency of a Signal-Part 2: Algorithms and Applications*. (1992).

12.	Rosenblum, M. G. *et al.* Locking-Based Frequency Measurement and Synchronization of Chaotic Oscillators with Complex Dynamics. *Phys Rev Lett* 89, (2002).

13.	Rosenblum, M., Pikovsky, A., Kühn, A. A. & Busch, J. L. Real-time estimation of phase and amplitude with application to neural data. *Sci Rep* 11, (2021).

14.	Schreglmann, S. R. *et al.* Non-invasive suppression of essential tremor via phase-locked disruption of its temporal coherence. *Nat Commun* 12, (2021).

15.	Zrenner, C. *et al.* The shaky ground truth of real-time phase estimation. *Neuroimage* 214, (2020).

16.	Gordon, P. C., Belardinelli, P., Stenroos, M., Ziemann, U. & Zrenner, C. Prefrontal theta phase-dependent rTMS-induced plasticity of cortical and behavioral responses in human cortex. *Brain Stimul* 15, 391–402 (2022).

17.	Hussain, S. J. *et al.* Phase-dependent offline enhancement of human motor memory. *Brain Stimul* 14, 873–883 (2021).

18.	Schaworonkow, N., Triesch, J., Ziemann, U. & Zrenner, C. EEG-triggered TMS reveals stronger brain state-dependent modulation of motor evoked potentials at weaker stimulation intensities. *Brain Stimul* 12, 110–118 (2019).

19.	Wodeyar, A., Schatza, M., Widge, A. S., Eden, U. T. & Kramer, M. A. A state space modeling approach to real-time phase estimation. (2021) doi:10.7554/eLife.

20.	McNamara, C. G., Rothwell, M. & Sharott, A. Stable, interactive modulation of neuronal oscillations produced through brain-machine equilibrium. *Cell Rep* 41, (2022).

21.	Busch, J. L., Feldmann, L. K., Kühn, A. A. & Rosenblum, M. Real-time phase and amplitude estimation of neurophysiological signals exploiting a non-resonant oscillator. *Exp Neurol* 347, (2022).
